# Variants in *ACE2* and *TMPRSS2* genes are not major determinants of COVID-19 severity in UK Biobank subjects

**DOI:** 10.1101/2020.05.01.20085860

**Authors:** David Curtis

## Abstract

It is plausible that variants in the *ACE2* and *TMPRSS2* genes might contribute to variation in COVID-19 severity and that these could explain why some people become very unwell whereas most do not. Exome sequence data was obtained for 49,953 UK Biobank subjects of whom 74 had tested positive for SARS-CoV-2 and could be presumed to have severe disease. A weighted burden analysis was carried out using SCOREASSOC to determine whether there were differences between these cases and the other sequenced subjects in the overall burden of rare, damaging variants in *ACE2* or *TMPRSS2*. There were no statistically significant differences in weighted burden scores between cases and controls for either gene. There were no individual DNA sequence variants with a markedly different frequency between cases and controls. Whether there are small effects on severity, or whether there might be rare variants with major effect sizes, would require studies in much larger samples. Genetic variants affecting the structure and function of the ACE2 and TMPRSS2 proteins are not a major determinant of whether infection with SARS-CoV-2 results in severe symptoms. This research has been conducted using the UK Biobank Resource.

## Introduction

There is wide variation in the severity of symptoms in patients infected with SARS-CoV-2 and there are reports in the UK that members of ethnic minorities are more severely affected. An obvious possible explanation for these findings would be that genetic polymorphisms affecting structure or function of key proteins could influence host susceptibility and/or responses to infection. If these polymorphisms varied in frequency between different ethnic groups then this could contribute to differential outcomes.

Two key proteins involved in SARS-CoV-2 infective processes are ACE2, which is expressed on the cell surface and acts as a receptor for the viral S protein, and TMPRSS2, which cleaves the S protein to allow fusion of the viral and cellular membranes (Hoffmann et al., 2020). Variants in the genes coding for these proteins might contribute to different responses to infection.

## Methods

The UK Biobank dataset was downloaded along with the variant call files for 49,953 subjects who had undergone exome-sequencing and genotyped using the GRCh38 assembly with coverage 20X at 94.6% of sites on average (Hout et al., 2019). Informed consent from the subjects and ethical approval for research uses of the data had been obtained by UK Biobank. All variants were annotated using VEP, PolyPhen and SIFT (Adzhubei et al., 2013; Kumar et al., 2009; McLaren et al., 2016). To obtain population principal components reflecting ancestry, version 1.90beta of *plink* (https://www.cog-genomics.org/plink2) was run with the options *--maf 0*.*1 --pca header tabs -- make-rel* (Chang et al., 2015; Purcell et al., 2007, 2009).

The COVID-19 results table was downloaded from UK Biobank on 28^th^ April 2020. This contained results for 2,724 subjects who had undergone testing for SARS-CoV-2 infection between 16^th^ March and 14^th^ April 2020 (Armstrong et al., 2020). During this period, testing in the UK was done almost exclusively on patients admitted to hospital and thus patients testing positive can be assumed to have severe disease because patients with milder symptoms were generally left at home. Of the subjects tested, 185 had been exome sequenced, of whom 74 had tested positive, meaning that they had at least one swab which demonstrated the presence of viral RNA at detectable levels. The proportion of infected subjects who require hospitalisation rises with age but is still only 0.18 for those aged 80 or over (Verity et al., 2020). Thus the subjects who tested positive could be regarded as cases with an unusually severe response to infection whereas the subjects who tested negative or who were not tested could be regarded as unscreened controls, most of whom would not have severe symptoms even if infected.

SCOREASSOC was then used to carry out a weighted burden analysis to test whether, in *ACE2* or *TMPRSS2*, sequence variants which were rarer and/or predicted to have more severe functional effects occurred more commonly in cases, i.e. subjects who tested positive for SARS-CoV-2, than all the other sequenced subjects. All available variants in each gene were included in the analyses. As originally described, variants were weighted according to frequency so that rare variants were accorded 10 times the weight of common variants (Curtis, 2012). Variants were additionally weighted according to their functional annotation using the default weights provided with the GENEVARASSOC program, which was used to generate input files for weighted burden analysis by SCOREASSOC (Curtis, 2019, 2016, 2012). For example, a weight of 5 was assigned for a synonymous variant, 10 for a non-synonymous variant and 20 for a stop gained variant. Additionally, 10 was added to the weight if the PolyPhen annotation was possibly or probably damaging and also if the SIFT annotation was deleterious, meaning that a non-synonymous variant annotated as both damaging and deleterious would be assigned an overall weight of 30. The full set of weights is shown in Table 1, copied from the previous reports which used this method (Curtis et al., 2019, 2018). Variants were excluded if there were more than 10% of genotypes missing in the controls or if the heterozygote count was smaller than both homozygote counts in the controls. For each variant, an overall weight was obtained consisting of the product of the frequency-based weight and the annotation-based weight. For each subject a gene-wise weighted burden score was derived as the sum of the variant-wise weights, each multiplied by the number of alleles of the variant which the given subject possessed. *ACE2* is located on the X chromosome and males were treated as if they were homozygotes. If a subject was not genotyped for a variant then they were assigned the subject-wise average score for that variant.

**Table 1.**
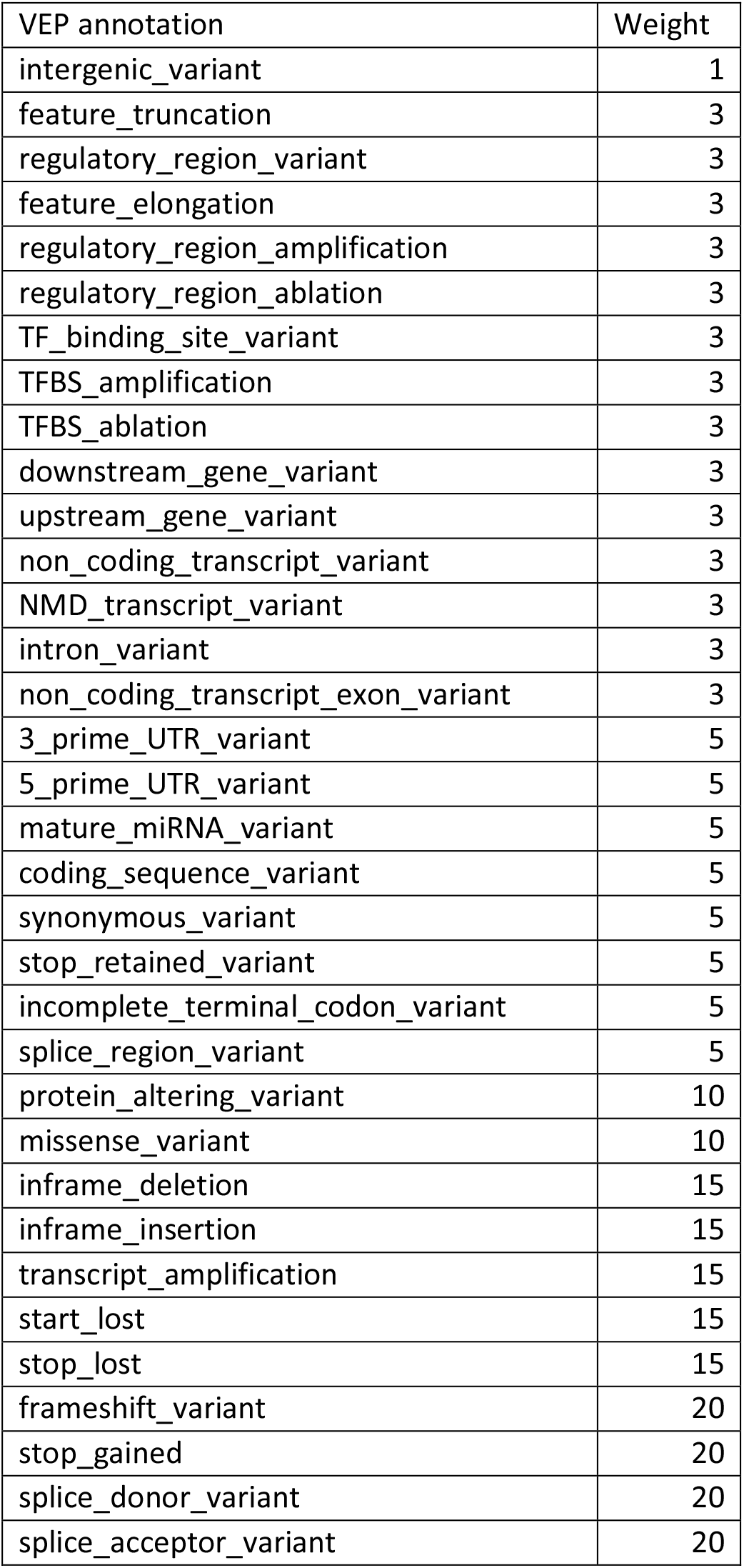
The table shows the weight accorded to each type of variant as annotated by VEP (McLaren et al., 2016). 10 was added to this weight if the variant was annotated by Polyphen as possibly or probably damaging and 10 was added if SIFT annotated it as deleterious (Adzhubei et al., 2013; Kumar et al., 2009).

A t test was carried out to determine whether the gene-wise burden scores differed between cases and controls and additionally ridge regression analysis with lamda=1 was performed incorporating the first 20 principal components, as described previously (Curtis, 2019; Curtis et al., 2019, 2018). To do this, SCOREASSOC first calculates the likelihood for the phenotypes as predicted by the principal components and then calculates the likelihood using a model which additionally incorporates the gene-wise burden scores. It then carries out a likelihood ratio test assuming that twice the natural log of the likelihood ratio follows a chi-squared distribution with one degree of freedom to produce a p value.

## Results

The genotype counts and frequencies of variants are presented in Supplementary Table 1, with variant positions and annotations redacted in order to preserve subject anonymity. There were 512 valid variants in *ACE2* and there was no tendency for the weighted burden scores to be different between cases (mean (sd): 25.9 (45.7)) and controls (22.6 (37.8)): t=0.74, 49951 df, p=0.45 and chi-squared=0.33, 1 df, p=0.57. There were 658 valid variants in *TMPRSS2* and although the weighted burden scores were lower in cases (64.6 (38.1)) than in controls (74.0 (48.9)) this difference would not meet conventional standards for statistical significance after applying a Bonferroni correction for the fact that two genes were tested: t=-1.6, 49951 df, p=0.10 and, in the ridge regression analysis incorporating principal components, chi-squared=4.17, 1 df, p = 0.04. On visual inspection of the results there were no individual variants with markedly different frequencies between cases and controls. Of course, for both genes there were many rare variants which were observed in controls but not in cases but this is as expected given the disparity in sample sizes.

## Discussion

Although the number of severely affected subjects who had been sequenced is very small it is nevertheless possible to draw some preliminary conclusions and given the importance of the topic it seems reasonable to communicate these findings. In general, the results are negative. It is not the case that a large proportion of severely affected subjects have a particular genetic variant in one of these genes which is relatively rare in the general population. Nor is it the case that there is a common variant which confers strong protection against severe infection. It remains possible that there might be rare variants which have a major effect on risk in individual subjects but such effects would only be detected with larger sample sizes.

The fact that the weighted burden scores were higher in controls than in cases is consistent with the hypothesis that rare genetic variants in *TMPRSS2* with functional effects disrupting functioning of the protein might be protective against severe infection. Although this is biologically plausible it should be emphasised that the results obtained are not statistically significant. This could be investigated further by carrying out targeted sequencing of this gene in a sample of a few hundred severely affected subjects.

Genetic variants affecting the structure and function of the ACE2 and TMPRSS2 proteins are not a major determinant of whether infection with SARS-CoV-2 results in severe symptoms.

## Data Availability

No new data was generated for this study. The data used is available on application from UK Biobank.

## Acknowledgments

This research has been conducted using the UK Biobank Resource. The author wishes to acknowledge the staff supporting the High Performance Computing Cluster, Computer Science Department, University College London. This work was carried out in part using resources provided by BBSRC equipment grant BB/R01356X/1.

---

**Supplementary Table 1** Table showing genotype counts and allele frequencies for variants in *ACE2* and *TMPRSS2* in cases tested positive for SARS-CoV-2, presumed to have severe illness, against background controls. Each variant is assigned a weight, with higher weights for variants which are rarer and/or with predicted damaging effects. For variants in *ACE2*, which is on the X chromosome, males are treated as homozygotes. To preserve subject anonymity, variant positions and annotations are redacted.

**Supplementary Table 1A.** Genotype counts and frequencies for variants in *ACE2*.

| Variant | contAA | contAB | contBB | contFreq | caseAA | caseAB | caseBB | caseFreq | Weight |
| --- | --- | --- | --- | --- | --- | --- | --- | --- | --- |
| 1 | 49866 | 1 | 0 | 0.00001 | 74 | 0 | 0 | 0 | 30 |
| 2 | 49020 | 472 | 236 | 0.009492 | 70 | 2 | 0 | 0.013889 | 28.98 |
| 3 | 49712 | 8 | 0 | 0.00008 | 74 | 0 | 0 | 0 | 29.99 |
| 4 | 49627 | 1 | 0 | 0.00001 | 73 | 0 | 0 | 0 | 30 |
| 5 | 49368 | 9 | 0 | 0.000091 | 73 | 0 | 0 | 0 | 29.99 |
| 6 | 49751 | 88 | 3 | 0.000943 | 74 | 0 | 0 | 0 | 29.9 |
| 7 | 49862 | 7 | 1 | 0.00009 | 74 | 0 | 0 | 0 | 29.99 |
| 8 | 49872 | 1 | 0 | 0.00001 | 74 | 0 | 0 | 0 | 30 |
| 9 | 49091 | 474 | 308 | 0.010928 | 70 | 2 | 2 | 0.040541 | 28.83 |
| 10 | 49877 | 2 | 0 | 0.00002 | 74 | 0 | 0 | 0 | 30 |
| 11 | 49878 | 1 | 0 | 0.00001 | 74 | 0 | 0 | 0 | 50 |
| 12 | 49878 | 1 | 0 | 0.00001 | 74 | 0 | 0 | 0 | 50 |
| 13 | 49876 | 2 | 1 | 0.00004 | 74 | 0 | 0 | 0 | 49.99 |
| 14 | 49871 | 6 | 1 | 0.00008 | 74 | 0 | 0 | 0 | 49.99 |
| 15 | 49878 | 1 | 0 | 0.00001 | 74 | 0 | 0 | 0 | 50 |
| 16 | 49867 | 10 | 2 | 0.00014 | 74 | 0 | 0 | 0 | 49.97 |
| 17 | 49878 | 1 | 0 | 0.00001 | 74 | 0 | 0 | 0 | 50 |
| 18 | 49878 | 1 | 0 | 0.00001 | 74 | 0 | 0 | 0 | 50 |
| 19 | 49877 | 1 | 1 | 0.00003 | 74 | 0 | 0 | 0 | 49.99 |
| 20 | 49878 | 1 | 0 | 0.00001 | 74 | 0 | 0 | 0 | 50 |
| 21 | 49878 | 1 | 0 | 0.00001 | 74 | 0 | 0 | 0 | 50 |
| 22 | 49878 | 1 | 0 | 0.00001 | 74 | 0 | 0 | 0 | 50 |
| 23 | 49878 | 1 | 0 | 0.00001 | 74 | 0 | 0 | 0 | 50 |
| 24 | 49854 | 2 | 0 | 0.00002 | 74 | 0 | 0 | 0 | 99.99 |
| 25 | 49870 | 7 | 2 | 0.00011 | 74 | 0 | 0 | 0 | 49.98 |
| 26 | 49822 | 37 | 19 | 0.000752 | 74 | 0 | 0 | 0 | 49.86 |
| 27 | 49874 | 3 | 2 | 0.00007 | 74 | 0 | 0 | 0 | 49.99 |
| 28 | 49873 | 3 | 3 | 0.00009 | 74 | 0 | 0 | 0 | 49.98 |
| 29 | 49878 | 1 | 0 | 0.00001 | 74 | 0 | 0 | 0 | 50 |
| 30 | 49878 | 1 | 0 | 0.00001 | 74 | 0 | 0 | 0 | 50 |
| 31 | 49875 | 2 | 2 | 0.00006 | 74 | 0 | 0 | 0 | 49.99 |
| 32 | 49877 | 2 | 0 | 0.00002 | 74 | 0 | 0 | 0 | 30 |
| 33 | 49876 | 2 | 1 | 0.00004 | 74 | 0 | 0 | 0 | 30 |
| 34 | 49868 | 2 | 0 | 0.00002 | 74 | 0 | 0 | 0 | 30 |
| 35 | 49854 | 4 | 0 | 0.00004 | 74 | 0 | 0 | 0 | 30 |
| 36 | 49505 | 1 | 0 | 0.00001 | 73 | 0 | 0 | 0 | 30 |
| 37 | 49628 | 1 | 0 | 0.00001 | 73 | 0 | 0 | 0 | 30 |
| 38 | 49468 | 120 | 7 | 0.001351 | 73 | 0 | 0 | 0 | 29.85 |
| 39 | 49737 | 1 | 0 | 0.00001 | 73 | 0 | 0 | 0 | 30 |
| 40 | 49819 | 4 | 0 | 0.00004 | 74 | 0 | 0 | 0 | 30 |
| 41 | 49877 | 1 | 1 | 0.00003 | 74 | 0 | 0 | 0 | 49.99 |
| 42 | 49878 | 1 | 0 | 0.00001 | 74 | 0 | 0 | 0 | 50 |
| 43 | 49878 | 1 | 0 | 0.00001 | 74 | 0 | 0 | 0 | 50 |
| 44 | 49871 | 3 | 0 | 0.00003 | 74 | 0 | 0 | 0 | 49.99 |
| 45 | 49868 | 1 | 0 | 0.00001 | 74 | 0 | 0 | 0 | 50 |
| 46 | 49838 | 1 | 0 | 0.00001 | 74 | 0 | 0 | 0 | 30 |
| 47 | 49833 | 1 | 0 | 0.00001 | 74 | 0 | 0 | 0 | 30 |
| 48 | 49831 | 1 | 0 | 0.00001 | 74 | 0 | 0 | 0 | 30 |
| 49 | 49770 | 32 | 5 | 0.000422 | 74 | 0 | 0 | 0 | 29.95 |
| 50 | 49684 | 11 | 0 | 0.000111 | 73 | 0 | 0 | 0 | 29.99 |
| 51 | 49655 | 2 | 0 | 0.00002 | 73 | 0 | 0 | 0 | 30 |
| 52 | 49635 | 3 | 0 | 0.00003 | 73 | 0 | 0 | 0 | 30 |
| 53 | 49589 | 1 | 0 | 0.00001 | 73 | 0 | 0 | 0 | 30 |
| 54 | 49278 | 2 | 0 | 0.00002 | 72 | 0 | 0 | 0 | 30 |
| 55 | 48863 | 2 | 0 | 0.00002 | 69 | 0 | 0 | 0 | 30 |
| 56 | 49281 | 2 | 0 | 0.00002 | 69 | 0 | 0 | 0 | 30 |
| 57 | 49670 | 5 | 0 | 0.00005 | 71 | 0 | 0 | 0 | 29.99 |
| 58 | 49700 | 2 | 0 | 0.00002 | 72 | 0 | 0 | 0 | 30 |
| 59 | 49818 | 11 | 0 | 0.00011 | 73 | 0 | 0 | 0 | 29.99 |
| 60 | 49136 | 440 | 217 | 0.008776 | 69 | 2 | 2 | 0.041096 | 29.06 |
| 61 | 49575 | 207 | 71 | 0.0035 | 74 | 0 | 0 | 0 | 49.37 |
| 62 | 49872 | 6 | 0 | 0.00006 | 74 | 0 | 0 | 0 | 49.99 |
| 63 | 49874 | 3 | 0 | 0.00003 | 74 | 0 | 0 | 0 | 49.99 |
| 64 | 49871 | 3 | 0 | 0.00003 | 74 | 0 | 0 | 0 | 49.99 |
| 65 | 49817 | 1 | 0 | 0.00001 | 74 | 0 | 0 | 0 | 30 |
| 66 | 49454 | 1 | 0 | 0.00001 | 73 | 0 | 0 | 0 | 30 |
| 67 | 49414 | 2 | 0 | 0.00002 | 73 | 0 | 0 | 0 | 30 |
| 68 | 49376 | 1 | 0 | 0.00001 | 73 | 0 | 0 | 0 | 30 |
| 69 | 48952 | 2 | 0 | 0.00002 | 73 | 0 | 0 | 0 | 30 |
| 70 | 49846 | 3 | 0 | 0.00003 | 74 | 0 | 0 | 0 | 30 |
| 71 | 49835 | 21 | 0 | 0.000211 | 74 | 0 | 0 | 0 | 29.98 |
| 72 | 49875 | 2 | 0 | 0.00002 | 74 | 0 | 0 | 0 | 50 |
| 73 | 49860 | 2 | 0 | 0.00002 | 74 | 0 | 0 | 0 | 30 |
| 74 | 48538 | 4 | 0 | 0.000041 | 68 | 0 | 0 | 0 | 30 |
| 75 | 49445 | 1 | 0 | 0.00001 | 74 | 0 | 0 | 0 | 30 |
| 76 | 49481 | 3 | 0 | 0.00003 | 74 | 0 | 0 | 0 | 30 |
| 77 | 49876 | 2 | 0 | 0.00002 | 74 | 0 | 0 | 0 | 50 |
| 78 | 49867 | 1 | 0 | 0.00001 | 74 | 0 | 0 | 0 | 50 |
| 79 | 49770 | 3 | 0 | 0.00003 | 74 | 0 | 0 | 0 | 30 |
| 80 | 49766 | 4 | 0 | 0.00004 | 74 | 0 | 0 | 0 | 30 |
| 81 | 49593 | 1 | 0 | 0.00001 | 74 | 0 | 0 | 0 | 30 |
| 82 | 49455 | 1 | 0 | 0.00001 | 74 | 0 | 0 | 0 | 30 |
| 83 | 49298 | 2 | 0 | 0.00002 | 74 | 0 | 0 | 0 | 30 |
| 84 | 48175 | 2 | 0 | 0.000021 | 69 | 0 | 0 | 0 | 30 |
| 85 | 48553 | 2 | 0 | 0.000021 | 72 | 0 | 0 | 0 | 30 |
| 86 | 49304 | 1 | 0 | 0.00001 | 72 | 0 | 0 | 0 | 30 |
| 87 | 49600 | 1 | 0 | 0.00001 | 73 | 0 | 0 | 0 | 30 |
| 88 | 49658 | 3 | 0 | 0.00003 | 73 | 0 | 0 | 0 | 30 |
| 89 | 49750 | 1 | 0 | 0.00001 | 74 | 0 | 0 | 0 | 50 |
| 90 | 49760 | 3 | 0 | 0.00003 | 74 | 0 | 0 | 0 | 199.98 |
| 91 | 49785 | 1 | 0 | 0.00001 | 74 | 0 | 0 | 0 | 50 |
| 92 | 49771 | 22 | 0 | 0.000221 | 74 | 0 | 0 | 0 | 199.84 |
| 93 | 49835 | 7 | 0 | 0.00007 | 74 | 0 | 0 | 0 | 49.99 |
| 94 | 49867 | 4 | 0 | 0.00004 | 74 | 0 | 0 | 0 | 49.99 |
| 95 | 49870 | 4 | 0 | 0.00004 | 74 | 0 | 0 | 0 | 49.99 |
| 96 | 49872 | 2 | 0 | 0.00002 | 74 | 0 | 0 | 0 | 50 |
| 97 | 49876 | 1 | 0 | 0.00001 | 74 | 0 | 0 | 0 | 149.99 |
| 98 | 49851 | 6 | 1 | 0.00008 | 74 | 0 | 0 | 0 | 29.99 |
| 99 | 49854 | 6 | 0 | 0.00006 | 74 | 0 | 0 | 0 | 29.99 |
| 100 | 49321 | 4 | 0 | 0.000041 | 74 | 0 | 0 | 0 | 30 |
| 101 | 48768 | 237 | 18 | 0.002784 | 74 | 0 | 0 | 0 | 29.7 |
| 102 | 49107 | 1 | 0 | 0.00001 | 74 | 0 | 0 | 0 | 30 |
| 103 | 49303 | 1 | 0 | 0.00001 | 73 | 0 | 0 | 0 | 30 |
| 104 | 49809 | 22 | 4 | 0.000301 | 74 | 0 | 0 | 0 | 29.97 |
| 105 | 49864 | 4 | 0 | 0.00004 | 74 | 0 | 0 | 0 | 30 |
| 106 | 49869 | 8 | 0 | 0.00008 | 74 | 0 | 0 | 0 | 49.99 |
| 107 | 49877 | 2 | 0 | 0.00002 | 74 | 0 | 0 | 0 | 50 |
| 108 | 49870 | 6 | 2 | 0.0001 | 74 | 0 | 0 | 0 | 49.98 |
| 109 | 49874 | 4 | 1 | 0.00006 | 74 | 0 | 0 | 0 | 49.99 |
| 110 | 49877 | 2 | 0 | 0.00002 | 73 | 1 | 0 | 0.006757 | 49.99 |
| 111 | 49793 | 1 | 0 | 0.00001 | 73 | 0 | 0 | 0 | 30 |
| 112 | 49643 | 1 | 0 | 0.00001 | 72 | 0 | 0 | 0 | 30 |
| 113 | 49390 | 8 | 0 | 0.000081 | 72 | 0 | 0 | 0 | 29.99 |
| 114 | 49254 | 16 | 0 | 0.000162 | 72 | 0 | 0 | 0 | 29.98 |
| 115 | 48174 | 10 | 0 | 0.000104 | 71 | 0 | 0 | 0 | 29.99 |
| 116 | 48059 | 43 | 0 | 0.000447 | 71 | 0 | 0 | 0 | 29.95 |
| 117 | 49584 | 2 | 0 | 0.00002 | 74 | 0 | 0 | 0 | 30 |
| 118 | 49615 | 37 | 2 | 0.000413 | 74 | 0 | 0 | 0 | 29.96 |
| 119 | 49841 | 1 | 0 | 0.00001 | 74 | 0 | 0 | 0 | 30 |
| 120 | 49877 | 2 | 0 | 0.00002 | 74 | 0 | 0 | 0 | 30 |
| 121 | 49867 | 10 | 2 | 0.00014 | 74 | 0 | 0 | 0 | 29.98 |
| 122 | 49876 | 3 | 0 | 0.00003 | 74 | 0 | 0 | 0 | 30 |
| 123 | 49878 | 1 | 0 | 0.00001 | 74 | 0 | 0 | 0 | 50 |
| 124 | 49623 | 187 | 69 | 0.003258 | 72 | 1 | 1 | 0.02027 | 49.41 |
| 125 | 49877 | 1 | 1 | 0.00003 | 74 | 0 | 0 | 0 | 49.99 |
| 126 | 49878 | 1 | 0 | 0.00001 | 74 | 0 | 0 | 0 | 50 |
| 127 | 49877 | 2 | 0 | 0.00002 | 74 | 0 | 0 | 0 | 30 |
| 128 | 49878 | 1 | 0 | 0.00001 | 74 | 0 | 0 | 0 | 30 |
| 129 | 48035 | 2 | 0 | 0.000021 | 72 | 0 | 0 | 0 | 30 |
| 130 | 48772 | 1 | 0 | 0.00001 | 72 | 0 | 0 | 0 | 30 |
| 131 | 48916 | 1 | 0 | 0.00001 | 72 | 0 | 0 | 0 | 30 |
| 132 | 49263 | 1 | 0 | 0.00001 | 74 | 0 | 0 | 0 | 30 |
| 133 | 49410 | 164 | 11 | 0.001876 | 72 | 1 | 0 | 0.006849 | 29.8 |
| 134 | 49818 | 2 | 0 | 0.00002 | 74 | 0 | 0 | 0 | 30 |
| 135 | 49857 | 1 | 0 | 0.00001 | 74 | 0 | 0 | 0 | 30 |
| 136 | 49874 | 3 | 1 | 0.00005 | 74 | 0 | 0 | 0 | 49.99 |
| 137 | 49876 | 2 | 0 | 0.00002 | 74 | 0 | 0 | 0 | 50 |
| 138 | 49850 | 14 | 10 | 0.000341 | 74 | 0 | 0 | 0 | 49.94 |
| 139 | 49877 | 1 | 1 | 0.00003 | 74 | 0 | 0 | 0 | 49.99 |
| 140 | 49878 | 1 | 0 | 0.00001 | 74 | 0 | 0 | 0 | 50 |
| 141 | 49874 | 3 | 1 | 0.00005 | 74 | 0 | 0 | 0 | 49.99 |
| 142 | 49878 | 1 | 0 | 0.00001 | 74 | 0 | 0 | 0 | 50 |
| 143 | 49878 | 1 | 0 | 0.00001 | 74 | 0 | 0 | 0 | 50 |
| 144 | 49878 | 1 | 0 | 0.00001 | 74 | 0 | 0 | 0 | 50 |
| 145 | 49878 | 1 | 0 | 0.00001 | 74 | 0 | 0 | 0 | 50 |
| 146 | 49878 | 1 | 0 | 0.00001 | 74 | 0 | 0 | 0 | 50 |
| 147 | 49875 | 3 | 1 | 0.00005 | 73 | 0 | 1 | 0.013514 | 49.99 |
| 148 | 49857 | 18 | 3 | 0.000241 | 74 | 0 | 0 | 0 | 49.96 |
| 149 | 49878 | 1 | 0 | 0.00001 | 74 | 0 | 0 | 0 | 30 |
| 150 | 49811 | 11 | 1 | 0.00013 | 73 | 1 | 0 | 0.006757 | 29.98 |
| 151 | 49481 | 5 | 0 | 0.000051 | 74 | 0 | 0 | 0 | 29.99 |
| 152 | 48991 | 4 | 0 | 0.000041 | 74 | 0 | 0 | 0 | 30 |
| 153 | 49280 | 17 | 0 | 0.000172 | 73 | 0 | 0 | 0 | 29.98 |
| 154 | 49366 | 1 | 0 | 0.00001 | 73 | 0 | 0 | 0 | 30 |
| 155 | 49775 | 2 | 0 | 0.00002 | 74 | 0 | 0 | 0 | 30 |
| 156 | 49868 | 1 | 0 | 0.00001 | 74 | 0 | 0 | 0 | 30 |
| 157 | 49873 | 1 | 0 | 0.00001 | 74 | 0 | 0 | 0 | 30 |
| 158 | 49877 | 2 | 0 | 0.00002 | 74 | 0 | 0 | 0 | 50 |
| 159 | 49857 | 6 | 0 | 0.00006 | 73 | 0 | 0 | 0 | 29.99 |
| 160 | 49858 | 5 | 0 | 0.00005 | 73 | 0 | 0 | 0 | 29.99 |
| 161 | 49856 | 2 | 0 | 0.00002 | 73 | 0 | 0 | 0 | 30 |
| 162 | 49538 | 3 | 0 | 0.00003 | 73 | 0 | 0 | 0 | 30 |
| 163 | 49504 | 1 | 0 | 0.00001 | 73 | 0 | 0 | 0 | 30 |
| 164 | 49490 | 1 | 0 | 0.00001 | 73 | 0 | 0 | 0 | 30 |
| 165 | 49368 | 25 | 2 | 0.000294 | 72 | 0 | 0 | 0 | 29.97 |
| 166 | 49288 | 3 | 0 | 0.00003 | 72 | 0 | 0 | 0 | 30 |
| 167 | 49289 | 3 | 0 | 0.00003 | 72 | 0 | 0 | 0 | 30 |
| 168 | 47532 | 2 | 0 | 0.000021 | 72 | 0 | 0 | 0 | 30 |
| 169 | 49702 | 3 | 0 | 0.00003 | 74 | 0 | 0 | 0 | 30 |
| 170 | 49877 | 2 | 0 | 0.00002 | 74 | 0 | 0 | 0 | 50 |
| 171 | 49873 | 4 | 2 | 0.00008 | 74 | 0 | 0 | 0 | 49.99 |
| 172 | 49877 | 1 | 1 | 0.00003 | 74 | 0 | 0 | 0 | 49.99 |
| 173 | 49877 | 2 | 0 | 0.00002 | 74 | 0 | 0 | 0 | 50 |
| 174 | 49878 | 1 | 0 | 0.00001 | 74 | 0 | 0 | 0 | 50 |
| 175 | 49868 | 3 | 0 | 0.00003 | 74 | 0 | 0 | 0 | 49.99 |
| 176 | 49794 | 4 | 0 | 0.00004 | 74 | 0 | 0 | 0 | 49.99 |
| 177 | 49617 | 2 | 0 | 0.00002 | 74 | 0 | 0 | 0 | 30 |
| 178 | 49509 | 1 | 0 | 0.00001 | 74 | 0 | 0 | 0 | 30 |
| 179 | 48694 | 3 | 0 | 0.000031 | 73 | 0 | 0 | 0 | 30 |
| 180 | 46768 | 1 | 0 | 0.000011 | 72 | 0 | 0 | 0 | 30 |
| 181 | 49845 | 3 | 0 | 0.00003 | 74 | 0 | 0 | 0 | 30 |
| 182 | 49877 | 2 | 0 | 0.00002 | 74 | 0 | 0 | 0 | 30 |
| 183 | 49878 | 1 | 0 | 0.00001 | 74 | 0 | 0 | 0 | 30 |
| 184 | 49878 | 1 | 0 | 0.00001 | 74 | 0 | 0 | 0 | 30 |
| 185 | 49878 | 1 | 0 | 0.00001 | 74 | 0 | 0 | 0 | 30 |
| 186 | 49877 | 2 | 0 | 0.00002 | 74 | 0 | 0 | 0 | 30 |
| 187 | 49878 | 1 | 0 | 0.00001 | 74 | 0 | 0 | 0 | 50 |
| 188 | 49877 | 1 | 1 | 0.00003 | 74 | 0 | 0 | 0 | 49.99 |
| 189 | 49878 | 1 | 0 | 0.00001 | 74 | 0 | 0 | 0 | 50 |
| 190 | 49878 | 1 | 0 | 0.00001 | 74 | 0 | 0 | 0 | 50 |
| 191 | 49878 | 1 | 0 | 0.00001 | 74 | 0 | 0 | 0 | 50 |
| 192 | 49876 | 2 | 1 | 0.00004 | 74 | 0 | 0 | 0 | 49.99 |
| 193 | 49878 | 1 | 0 | 0.00001 | 74 | 0 | 0 | 0 | 50 |
| 194 | 49877 | 2 | 0 | 0.00002 | 74 | 0 | 0 | 0 | 50 |
| 195 | 49875 | 4 | 0 | 0.00004 | 74 | 0 | 0 | 0 | 49.99 |
| 196 | 49878 | 1 | 0 | 0.00001 | 74 | 0 | 0 | 0 | 50 |
| 197 | 49878 | 1 | 0 | 0.00001 | 74 | 0 | 0 | 0 | 50 |
| 198 | 49876 | 3 | 0 | 0.00003 | 74 | 0 | 0 | 0 | 49.99 |
| 199 | 49877 | 1 | 1 | 0.00003 | 74 | 0 | 0 | 0 | 199.98 |
| 200 | 49878 | 1 | 0 | 0.00001 | 74 | 0 | 0 | 0 | 50 |
| 201 | 49877 | 1 | 1 | 0.00003 | 74 | 0 | 0 | 0 | 30 |
| 202 | 49877 | 2 | 0 | 0.00002 | 74 | 0 | 0 | 0 | 30 |
| 203 | 49853 | 10 | 1 | 0.00012 | 74 | 0 | 0 | 0 | 29.99 |
| 204 | 49858 | 2 | 0 | 0.00002 | 74 | 0 | 0 | 0 | 30 |
| 205 | 49837 | 1 | 0 | 0.00001 | 74 | 0 | 0 | 0 | 30 |
| 206 | 49764 | 33 | 0 | 0.000331 | 73 | 1 | 0 | 0.006757 | 29.96 |
| 207 | 49798 | 1 | 0 | 0.00001 | 74 | 0 | 0 | 0 | 30 |
| 208 | 49644 | 112 | 11 | 0.001346 | 74 | 0 | 0 | 0 | 29.86 |
| 209 | 49798 | 3 | 0 | 0.00003 | 74 | 0 | 0 | 0 | 30 |
| 210 | 49155 | 1 | 0 | 0.00001 | 71 | 0 | 0 | 0 | 30 |
| 211 | 49877 | 2 | 0 | 0.00002 | 74 | 0 | 0 | 0 | 50 |
| 212 | 49878 | 1 | 0 | 0.00001 | 74 | 0 | 0 | 0 | 50 |
| 213 | 49874 | 3 | 2 | 0.00007 | 74 | 0 | 0 | 0 | 49.99 |
| 214 | 49878 | 1 | 0 | 0.00001 | 74 | 0 | 0 | 0 | 50 |
| 215 | 49878 | 1 | 0 | 0.00001 | 74 | 0 | 0 | 0 | 50 |
| 216 | 49876 | 2 | 1 | 0.00004 | 74 | 0 | 0 | 0 | 49.99 |
| 217 | 49872 | 5 | 0 | 0.00005 | 74 | 0 | 0 | 0 | 49.99 |
| 218 | 49786 | 4 | 0 | 0.00004 | 74 | 0 | 0 | 0 | 49.99 |
| 219 | 49323 | 2 | 0 | 0.00002 | 74 | 0 | 0 | 0 | 50 |
| 220 | 48576 | 2 | 0 | 0.000021 | 72 | 0 | 0 | 0 | 50 |
| 221 | 49812 | 2 | 0 | 0.00002 | 74 | 0 | 0 | 0 | 50 |
| 222 | 49827 | 1 | 0 | 0.00001 | 74 | 0 | 0 | 0 | 50 |
| 223 | 49854 | 3 | 0 | 0.00003 | 74 | 0 | 0 | 0 | 49.99 |
| 224 | 49864 | 1 | 0 | 0.00001 | 74 | 0 | 0 | 0 | 50 |
| 225 | 49878 | 1 | 0 | 0.00001 | 74 | 0 | 0 | 0 | 50 |
| 226 | 49875 | 4 | 0 | 0.00004 | 74 | 0 | 0 | 0 | 49.99 |
| 227 | 49878 | 1 | 0 | 0.00001 | 74 | 0 | 0 | 0 | 50 |
| 228 | 49874 | 5 | 0 | 0.00005 | 74 | 0 | 0 | 0 | 49.99 |
| 229 | 49878 | 1 | 0 | 0.00001 | 74 | 0 | 0 | 0 | 50 |
| 230 | 49876 | 2 | 0 | 0.00002 | 74 | 0 | 0 | 0 | 30 |
| 231 | 49874 | 1 | 0 | 0.00001 | 74 | 0 | 0 | 0 | 30 |
| 232 | 49868 | 2 | 0 | 0.00002 | 74 | 0 | 0 | 0 | 30 |
| 233 | 49813 | 8 | 0 | 0.00008 | 74 | 0 | 0 | 0 | 29.99 |
| 234 | 49719 | 1 | 0 | 0.00001 | 74 | 0 | 0 | 0 | 30 |
| 235 | 49695 | 2 | 0 | 0.00002 | 74 | 0 | 0 | 0 | 30 |
| 236 | 49766 | 3 | 0 | 0.00003 | 73 | 0 | 0 | 0 | 30 |
| 237 | 46504 | 2267 | 1107 | 0.04492 | 73 | 0 | 1 | 0.013514 | 42.29 |
| 238 | 49862 | 13 | 4 | 0.000211 | 74 | 0 | 0 | 0 | 49.96 |
| 239 | 49848 | 28 | 3 | 0.000341 | 74 | 0 | 0 | 0 | 49.94 |
| 240 | 47866 | 1396 | 617 | 0.026364 | 73 | 0 | 1 | 0.013514 | 45.38 |
| 241 | 49869 | 6 | 4 | 0.00014 | 74 | 0 | 0 | 0 | 49.97 |
| 242 | 49870 | 7 | 1 | 0.00009 | 74 | 0 | 0 | 0 | 49.98 |
| 243 | 49878 | 1 | 0 | 0.00001 | 74 | 0 | 0 | 0 | 50 |
| 244 | 49876 | 3 | 0 | 0.00003 | 74 | 0 | 0 | 0 | 49.99 |
| 245 | 49741 | 5 | 0 | 0.00005 | 74 | 0 | 0 | 0 | 29.99 |
| 246 | 48903 | 80 | 0 | 0.000817 | 71 | 0 | 0 | 0 | 29.91 |
| 247 | 49570 | 22 | 1 | 0.000242 | 72 | 0 | 0 | 0 | 29.97 |
| 248 | 49653 | 8 | 0 | 0.000081 | 72 | 0 | 0 | 0 | 29.99 |
| 249 | 49842 | 2 | 0 | 0.00002 | 74 | 0 | 0 | 0 | 30 |
| 250 | 49878 | 1 | 0 | 0.00001 | 74 | 0 | 0 | 0 | 30 |
| 251 | 49877 | 2 | 0 | 0.00002 | 74 | 0 | 0 | 0 | 30 |
| 252 | 49877 | 2 | 0 | 0.00002 | 74 | 0 | 0 | 0 | 30 |
| 253 | 49877 | 2 | 0 | 0.00002 | 74 | 0 | 0 | 0 | 30 |
| 254 | 49871 | 6 | 2 | 0.0001 | 74 | 0 | 0 | 0 | 49.98 |
| 255 | 49874 | 4 | 1 | 0.00006 | 74 | 0 | 0 | 0 | 49.99 |
| 256 | 49868 | 8 | 3 | 0.00014 | 74 | 0 | 0 | 0 | 49.97 |
| 257 | 49718 | 114 | 47 | 0.002085 | 74 | 0 | 0 | 0 | 49.63 |
| 258 | 49875 | 3 | 1 | 0.00005 | 74 | 0 | 0 | 0 | 49.99 |
| 259 | 49878 | 1 | 0 | 0.00001 | 74 | 0 | 0 | 0 | 50 |
| 260 | 49876 | 3 | 0 | 0.00003 | 74 | 0 | 0 | 0 | 149.98 |
| 261 | 49878 | 1 | 0 | 0.00001 | 74 | 0 | 0 | 0 | 50 |
| 262 | 49877 | 1 | 0 | 0.00001 | 74 | 0 | 0 | 0 | 50 |
| 263 | 49679 | 1 | 0 | 0.00001 | 74 | 0 | 0 | 0 | 30 |
| 264 | 49300 | 1 | 0 | 0.00001 | 73 | 0 | 0 | 0 | 30 |
| 265 | 49798 | 1 | 0 | 0.00001 | 74 | 0 | 0 | 0 | 30 |
| 266 | 49808 | 1 | 0 | 0.00001 | 74 | 0 | 0 | 0 | 30 |
| 267 | 49872 | 1 | 0 | 0.00001 | 74 | 0 | 0 | 0 | 30 |
| 268 | 49877 | 1 | 0 | 0.00001 | 74 | 0 | 0 | 0 | 30 |
| 269 | 49873 | 5 | 0 | 0.00005 | 74 | 0 | 0 | 0 | 29.99 |
| 270 | 49876 | 3 | 0 | 0.00003 | 74 | 0 | 0 | 0 | 30 |
| 271 | 49877 | 1 | 1 | 0.00003 | 74 | 0 | 0 | 0 | 30 |
| 272 | 49878 | 1 | 0 | 0.00001 | 74 | 0 | 0 | 0 | 30 |
| 273 | 49878 | 1 | 0 | 0.00001 | 74 | 0 | 0 | 0 | 50 |
| 274 | 49878 | 1 | 0 | 0.00001 | 74 | 0 | 0 | 0 | 50 |
| 275 | 49877 | 1 | 1 | 0.00003 | 74 | 0 | 0 | 0 | 49.99 |
| 276 | 49875 | 3 | 1 | 0.00005 | 73 | 0 | 1 | 0.013514 | 49.99 |
| 277 | 49878 | 1 | 0 | 0.00001 | 74 | 0 | 0 | 0 | 30 |
| 278 | 49843 | 18 | 2 | 0.000221 | 74 | 0 | 0 | 0 | 29.98 |
| 279 | 49852 | 1 | 0 | 0.00001 | 74 | 0 | 0 | 0 | 30 |
| 280 | 49850 | 1 | 0 | 0.00001 | 74 | 0 | 0 | 0 | 30 |
| 281 | 49814 | 2 | 0 | 0.00002 | 74 | 0 | 0 | 0 | 30 |
| 282 | 49794 | 2 | 0 | 0.00002 | 74 | 0 | 0 | 0 | 30 |
| 283 | 49792 | 4 | 0 | 0.00004 | 74 | 0 | 0 | 0 | 30 |
| 284 | 49800 | 2 | 0 | 0.00002 | 74 | 0 | 0 | 0 | 30 |
| 285 | 49803 | 1 | 0 | 0.00001 | 74 | 0 | 0 | 0 | 30 |
| 286 | 49848 | 1 | 0 | 0.00001 | 74 | 0 | 0 | 0 | 30 |
| 287 | 49853 | 3 | 0 | 0.00003 | 74 | 0 | 0 | 0 | 30 |
| 288 | 49875 | 2 | 0 | 0.00002 | 74 | 0 | 0 | 0 | 30 |
| 289 | 49872 | 6 | 1 | 0.00008 | 74 | 0 | 0 | 0 | 29.99 |
| 290 | 49878 | 1 | 0 | 0.00001 | 74 | 0 | 0 | 0 | 50 |
| 291 | 49843 | 1 | 0 | 0.00001 | 74 | 0 | 0 | 0 | 100 |
| 292 | 49852 | 9 | 0 | 0.00009 | 74 | 0 | 0 | 0 | 29.99 |
| 293 | 49726 | 7 | 1 | 0.00009 | 73 | 0 | 0 | 0 | 29.99 |
| 294 | 48489 | 1 | 0 | 0.00001 | 69 | 0 | 0 | 0 | 30 |
| 295 | 49365 | 45 | 0 | 0.000455 | 73 | 0 | 0 | 0 | 29.95 |
| 296 | 49465 | 5 | 0 | 0.000051 | 73 | 0 | 0 | 0 | 29.99 |
| 297 | 49778 | 8 | 0 | 0.00008 | 74 | 0 | 0 | 0 | 29.99 |
| 298 | 49762 | 28 | 0 | 0.000281 | 74 | 0 | 0 | 0 | 29.97 |
| 299 | 49601 | 204 | 49 | 0.003029 | 73 | 0 | 0 | 0 | 29.67 |
| 300 | 49872 | 2 | 0 | 0.00002 | 74 | 0 | 0 | 0 | 30 |
| 301 | 49871 | 6 | 0 | 0.00006 | 74 | 0 | 0 | 0 | 29.99 |
| 302 | 0 | 1 | 49855 | 0.99999 | 0 | 0 | 74 | 1 | 50 |
| 303 | 49855 | 1 | 0 | 0.00001 | 74 | 0 | 0 | 0 | 50 |
| 304 | 49876 | 2 | 0 | 0.00002 | 74 | 0 | 0 | 0 | 50 |
| 305 | 49876 | 3 | 0 | 0.00003 | 74 | 0 | 0 | 0 | 49.99 |
| 306 | 49877 | 2 | 0 | 0.00002 | 74 | 0 | 0 | 0 | 50 |
| 307 | 49876 | 2 | 1 | 0.00004 | 74 | 0 | 0 | 0 | 49.99 |
| 308 | 49878 | 1 | 0 | 0.00001 | 74 | 0 | 0 | 0 | 50 |
| 309 | 49877 | 2 | 0 | 0.00002 | 74 | 0 | 0 | 0 | 50 |
| 310 | 49836 | 26 | 15 | 0.000561 | 74 | 0 | 0 | 0 | 49.9 |
| 311 | 49864 | 11 | 3 | 0.00017 | 74 | 0 | 0 | 0 | 29.98 |
| 312 | 49868 | 3 | 0 | 0.00003 | 74 | 0 | 0 | 0 | 30 |
| 313 | 49858 | 1 | 0 | 0.00001 | 74 | 0 | 0 | 0 | 30 |
| 314 | 49788 | 1 | 0 | 0.00001 | 73 | 0 | 0 | 0 | 30 |
| 315 | 49606 | 112 | 15 | 0.001428 | 72 | 0 | 0 | 0 | 29.85 |
| 316 | 49569 | 29 | 1 | 0.000313 | 71 | 1 | 0 | 0.006944 | 29.97 |
| 317 | 49679 | 7 | 0 | 0.00007 | 74 | 0 | 0 | 0 | 29.99 |
| 318 | 49794 | 2 | 0 | 0.00002 | 74 | 0 | 0 | 0 | 30 |
| 319 | 49800 | 2 | 1 | 0.00004 | 74 | 0 | 0 | 0 | 30 |
| 320 | 49878 | 1 | 0 | 0.00001 | 74 | 0 | 0 | 0 | 50 |
| 321 | 49876 | 2 | 0 | 0.00002 | 74 | 0 | 0 | 0 | 50 |
| 322 | 49877 | 2 | 0 | 0.00002 | 74 | 0 | 0 | 0 | 50 |
| 323 | 49857 | 8 | 4 | 0.00016 | 74 | 0 | 0 | 0 | 29.98 |
| 324 | 49863 | 5 | 1 | 0.00007 | 74 | 0 | 0 | 0 | 29.99 |
| 325 | 49863 | 5 | 0 | 0.00005 | 74 | 0 | 0 | 0 | 29.99 |
| 326 | 49858 | 6 | 0 | 0.00006 | 74 | 0 | 0 | 0 | 29.99 |
| 327 | 49850 | 6 | 0 | 0.00006 | 73 | 0 | 0 | 0 | 29.99 |
| 328 | 49669 | 1 | 0 | 0.00001 | 73 | 0 | 0 | 0 | 30 |
| 329 | 49605 | 1 | 0 | 0.00001 | 73 | 0 | 0 | 0 | 30 |
| 330 | 49307 | 8 | 0 | 0.000081 | 71 | 0 | 0 | 0 | 29.99 |
| 331 | 49295 | 2 | 0 | 0.00002 | 71 | 0 | 0 | 0 | 30 |
| 332 | 48301 | 1 | 0 | 0.00001 | 69 | 0 | 0 | 0 | 30 |
| 333 | 49615 | 7 | 1 | 0.000091 | 73 | 0 | 0 | 0 | 29.99 |
| 334 | 49826 | 3 | 0 | 0.00003 | 73 | 0 | 0 | 0 | 30 |
| 335 | 49874 | 1 | 0 | 0.00001 | 74 | 0 | 0 | 0 | 50 |
| 336 | 49870 | 8 | 0 | 0.00008 | 74 | 0 | 0 | 0 | 49.99 |
| 337 | 49877 | 1 | 0 | 0.00001 | 74 | 0 | 0 | 0 | 50 |
| 338 | 49872 | 4 | 1 | 0.00006 | 74 | 0 | 0 | 0 | 49.99 |
| 339 | 49841 | 3 | 0 | 0.00003 | 74 | 0 | 0 | 0 | 49.99 |
| 340 | 49839 | 1 | 0 | 0.00001 | 74 | 0 | 0 | 0 | 50 |
| 341 | 49551 | 1 | 0 | 0.00001 | 73 | 0 | 0 | 0 | 30 |
| 342 | 48564 | 8 | 0 | 0.000082 | 72 | 0 | 0 | 0 | 29.99 |
| 343 | 48182 | 1 | 0 | 0.00001 | 71 | 0 | 0 | 0 | 30 |
| 344 | 47885 | 1 | 0 | 0.00001 | 71 | 0 | 0 | 0 | 30 |
| 345 | 47288 | 134 | 0 | 0.001413 | 71 | 0 | 0 | 0 | 29.85 |
| 346 | 47226 | 5 | 0 | 0.000053 | 70 | 0 | 0 | 0 | 29.99 |
| 347 | 0 | 1 | 47226 | 0.999989 | 0 | 0 | 70 | 1 | 30 |
| 348 | 0 | 10 | 47838 | 0.999896 | 0 | 0 | 73 | 1 | 29.99 |
| 349 | 47838 | 4 | 0 | 0.000042 | 73 | 0 | 0 | 0 | 30 |
| 350 | 49556 | 1 | 0 | 0.00001 | 73 | 0 | 0 | 0 | 30 |
| 351 | 49663 | 2 | 0 | 0.00002 | 74 | 0 | 0 | 0 | 30 |
| 352 | 49742 | 1 | 0 | 0.00001 | 74 | 0 | 0 | 0 | 30 |
| 353 | 49817 | 1 | 0 | 0.00001 | 74 | 0 | 0 | 0 | 30 |
| 354 | 49841 | 1 | 0 | 0.00001 | 74 | 0 | 0 | 0 | 30 |
| 355 | 49864 | 1 | 0 | 0.00001 | 74 | 0 | 0 | 0 | 30 |
| 356 | 49878 | 1 | 0 | 0.00001 | 74 | 0 | 0 | 0 | 50 |
| 357 | 49873 | 4 | 1 | 0.00006 | 74 | 0 | 0 | 0 | 49.99 |
| 358 | 49877 | 2 | 0 | 0.00002 | 74 | 0 | 0 | 0 | 50 |
| 359 | 49878 | 1 | 0 | 0.00001 | 74 | 0 | 0 | 0 | 50 |
| 360 | 49876 | 3 | 0 | 0.00003 | 74 | 0 | 0 | 0 | 49.99 |
| 361 | 49875 | 4 | 0 | 0.00004 | 74 | 0 | 0 | 0 | 49.99 |
| 362 | 49878 | 1 | 0 | 0.00001 | 74 | 0 | 0 | 0 | 50 |
| 363 | 49872 | 2 | 0 | 0.00002 | 74 | 0 | 0 | 0 | 50 |
| 364 | 49848 | 1 | 0 | 0.00001 | 74 | 0 | 0 | 0 | 30 |
| 365 | 49844 | 1 | 0 | 0.00001 | 74 | 0 | 0 | 0 | 30 |
| 366 | 49760 | 5 | 0 | 0.00005 | 74 | 0 | 0 | 0 | 29.99 |
| 367 | 49751 | 4 | 0 | 0.00004 | 74 | 0 | 0 | 0 | 30 |
| 368 | 49679 | 1 | 0 | 0.00001 | 73 | 0 | 0 | 0 | 30 |
| 369 | 49606 | 1 | 0 | 0.00001 | 73 | 0 | 0 | 0 | 30 |
| 370 | 49404 | 39 | 0 | 0.000394 | 74 | 0 | 0 | 0 | 29.96 |
| 371 | 49698 | 1 | 0 | 0.00001 | 74 | 0 | 0 | 0 | 30 |
| 372 | 49873 | 4 | 0 | 0.00004 | 74 | 0 | 0 | 0 | 49.99 |
| 373 | 49877 | 2 | 0 | 0.00002 | 74 | 0 | 0 | 0 | 50 |
| 374 | 49877 | 2 | 0 | 0.00002 | 74 | 0 | 0 | 0 | 50 |
| 375 | 49878 | 1 | 0 | 0.00001 | 74 | 0 | 0 | 0 | 50 |
| 376 | 49875 | 4 | 0 | 0.00004 | 74 | 0 | 0 | 0 | 49.99 |
| 377 | 49878 | 1 | 0 | 0.00001 | 74 | 0 | 0 | 0 | 50 |
| 378 | 49877 | 1 | 0 | 0.00001 | 74 | 0 | 0 | 0 | 50 |
| 379 | 49860 | 11 | 6 | 0.000231 | 74 | 0 | 0 | 0 | 49.96 |
| 380 | 49871 | 1 | 0 | 0.00001 | 74 | 0 | 0 | 0 | 50 |
| 381 | 49817 | 1 | 0 | 0.00001 | 74 | 0 | 0 | 0 | 30 |
| 382 | 48495 | 1 | 0 | 0.00001 | 71 | 0 | 0 | 0 | 30 |
| 383 | 48427 | 2 | 0 | 0.000021 | 69 | 0 | 0 | 0 | 30 |
| 384 | 49875 | 1 | 0 | 0.00001 | 74 | 0 | 0 | 0 | 30 |
| 385 | 49875 | 4 | 0 | 0.00004 | 74 | 0 | 0 | 0 | 49.99 |
| 386 | 49877 | 2 | 0 | 0.00002 | 74 | 0 | 0 | 0 | 50 |
| 387 | 49858 | 15 | 5 | 0.000251 | 74 | 0 | 0 | 0 | 49.95 |
| 388 | 49870 | 7 | 2 | 0.00011 | 74 | 0 | 0 | 0 | 49.98 |
| 389 | 49875 | 4 | 0 | 0.00004 | 74 | 0 | 0 | 0 | 49.99 |
| 390 | 49878 | 1 | 0 | 0.00001 | 74 | 0 | 0 | 0 | 50 |
| 391 | 49875 | 1 | 0 | 0.00001 | 74 | 0 | 0 | 0 | 50 |
| 392 | 49873 | 2 | 0 | 0.00002 | 74 | 0 | 0 | 0 | 30 |
| 393 | 49308 | 11 | 0 | 0.000112 | 73 | 0 | 0 | 0 | 29.99 |
| 394 | 49287 | 5 | 0 | 0.000051 | 73 | 0 | 0 | 0 | 29.99 |
| 395 | 49239 | 12 | 0 | 0.000122 | 73 | 0 | 0 | 0 | 29.99 |
| 396 | 49047 | 7 | 1 | 0.000092 | 72 | 0 | 0 | 0 | 29.99 |
| 397 | 49054 | 1 | 0 | 0.00001 | 72 | 0 | 0 | 0 | 30 |
| 398 | 49023 | 2 | 0 | 0.00002 | 72 | 0 | 0 | 0 | 30 |
| 399 | 49694 | 2 | 0 | 0.00002 | 73 | 0 | 0 | 0 | 30 |
| 400 | 48882 | 582 | 74 | 0.007368 | 72 | 1 | 0 | 0.006849 | 29.21 |
| 401 | 49706 | 1 | 0 | 0.00001 | 73 | 0 | 0 | 0 | 30 |
| 402 | 49761 | 2 | 0 | 0.00002 | 74 | 0 | 0 | 0 | 30 |
| 403 | 49877 | 2 | 0 | 0.00002 | 74 | 0 | 0 | 0 | 50 |
| 404 | 49878 | 1 | 0 | 0.00001 | 74 | 0 | 0 | 0 | 50 |
| 405 | 49878 | 1 | 0 | 0.00001 | 74 | 0 | 0 | 0 | 50 |
| 406 | 49875 | 4 | 0 | 0.00004 | 74 | 0 | 0 | 0 | 49.99 |
| 407 | 49869 | 8 | 1 | 0.0001 | 74 | 0 | 0 | 0 | 49.98 |
| 408 | 49878 | 1 | 0 | 0.00001 | 74 | 0 | 0 | 0 | 50 |
| 409 | 49858 | 1 | 0 | 0.00001 | 74 | 0 | 0 | 0 | 30 |
| 410 | 49834 | 1 | 1 | 0.00003 | 74 | 0 | 0 | 0 | 30 |
| 411 | 49830 | 1 | 0 | 0.00001 | 74 | 0 | 0 | 0 | 30 |
| 412 | 49830 | 1 | 0 | 0.00001 | 74 | 0 | 0 | 0 | 30 |
| 413 | 49823 | 1 | 0 | 0.00001 | 74 | 0 | 0 | 0 | 30 |
| 414 | 49642 | 1 | 0 | 0.00001 | 74 | 0 | 0 | 0 | 30 |
| 415 | 49605 | 1 | 0 | 0.00001 | 74 | 0 | 0 | 0 | 30 |
| 416 | 49214 | 11 | 0 | 0.000112 | 74 | 0 | 0 | 0 | 29.99 |
| 417 | 49525 | 2 | 0 | 0.00002 | 74 | 0 | 0 | 0 | 30 |
| 418 | 49580 | 6 | 0 | 0.000061 | 74 | 0 | 0 | 0 | 29.99 |
| 419 | 49871 | 2 | 0 | 0.00002 | 74 | 0 | 0 | 0 | 30 |
| 420 | 49457 | 289 | 117 | 0.005244 | 71 | 1 | 2 | 0.033784 | 29.43 |
| 421 | 49853 | 20 | 2 | 0.000241 | 74 | 0 | 0 | 0 | 29.97 |
| 422 | 49868 | 9 | 1 | 0.00011 | 74 | 0 | 0 | 0 | 29.99 |
| 423 | 49878 | 1 | 0 | 0.00001 | 74 | 0 | 0 | 0 | 50 |
| 424 | 49847 | 22 | 9 | 0.000401 | 74 | 0 | 0 | 0 | 49.93 |
| 425 | 49878 | 1 | 0 | 0.00001 | 74 | 0 | 0 | 0 | 199.99 |
| 426 | 49878 | 1 | 0 | 0.00001 | 74 | 0 | 0 | 0 | 50 |
| 427 | 49586 | 2 | 0 | 0.00002 | 74 | 0 | 0 | 0 | 30 |
| 428 | 49606 | 1 | 0 | 0.00001 | 74 | 0 | 0 | 0 | 30 |
| 429 | 49861 | 4 | 2 | 0.00008 | 74 | 0 | 0 | 0 | 29.99 |
| 430 | 49876 | 1 | 0 | 0.00001 | 74 | 0 | 0 | 0 | 30 |
| 431 | 49873 | 4 | 1 | 0.00006 | 74 | 0 | 0 | 0 | 49.99 |
| 432 | 49877 | 1 | 1 | 0.00003 | 74 | 0 | 0 | 0 | 49.99 |
| 433 | 49878 | 1 | 0 | 0.00001 | 74 | 0 | 0 | 0 | 50 |
| 434 | 49873 | 6 | 0 | 0.00006 | 74 | 0 | 0 | 0 | 49.99 |
| 435 | 49795 | 63 | 21 | 0.001053 | 74 | 0 | 0 | 0 | 49.81 |
| 436 | 49877 | 1 | 1 | 0.00003 | 74 | 0 | 0 | 0 | 49.99 |
| 437 | 49646 | 167 | 66 | 0.002997 | 74 | 0 | 0 | 0 | 49.46 |
| 438 | 49877 | 2 | 0 | 0.00002 | 74 | 0 | 0 | 0 | 50 |
| 439 | 49799 | 54 | 26 | 0.001063 | 74 | 0 | 0 | 0 | 49.81 |
| 440 | 49872 | 3 | 1 | 0.00005 | 74 | 0 | 0 | 0 | 199.96 |
| 441 | 49872 | 3 | 0 | 0.00003 | 74 | 0 | 0 | 0 | 49.99 |
| 442 | 49857 | 18 | 1 | 0.0002 | 74 | 0 | 0 | 0 | 29.98 |
| 443 | 49434 | 443 | 0 | 0.004441 | 72 | 2 | 0 | 0.013514 | 29.52 |
| 444 | 49874 | 1 | 0 | 0.00001 | 74 | 0 | 0 | 0 | 30 |
| 445 | 49859 | 1 | 0 | 0.00001 | 74 | 0 | 0 | 0 | 30 |
| 446 | 49809 | 1 | 0 | 0.00001 | 74 | 0 | 0 | 0 | 30 |
| 447 | 49737 | 1 | 0 | 0.00001 | 74 | 0 | 0 | 0 | 30 |
| 448 | 49737 | 1 | 0 | 0.00001 | 74 | 0 | 0 | 0 | 30 |
| 449 | 49834 | 21 | 5 | 0.000311 | 74 | 0 | 0 | 0 | 29.97 |
| 450 | 49872 | 5 | 0 | 0.00005 | 74 | 0 | 0 | 0 | 29.99 |
| 451 | 49878 | 1 | 0 | 0.00001 | 74 | 0 | 0 | 0 | 50 |
| 452 | 49878 | 1 | 0 | 0.00001 | 74 | 0 | 0 | 0 | 50 |
| 453 | 49876 | 3 | 0 | 0.00003 | 74 | 0 | 0 | 0 | 49.99 |
| 454 | 49873 | 4 | 1 | 0.00006 | 74 | 0 | 0 | 0 | 49.99 |
| 455 | 49878 | 1 | 0 | 0.00001 | 74 | 0 | 0 | 0 | 50 |
| 456 | 49878 | 1 | 0 | 0.00001 | 74 | 0 | 0 | 0 | 50 |
| 457 | 49857 | 5 | 0 | 0.00005 | 74 | 0 | 0 | 0 | 29.99 |
| 458 | 49748 | 21 | 0 | 0.000211 | 74 | 0 | 0 | 0 | 29.98 |
| 459 | 49708 | 6 | 0 | 0.00006 | 74 | 0 | 0 | 0 | 29.99 |
| 460 | 49704 | 6 | 0 | 0.00006 | 74 | 0 | 0 | 0 | 29.99 |
| 461 | 49705 | 6 | 0 | 0.00006 | 74 | 0 | 0 | 0 | 29.99 |
| 462 | 49693 | 6 | 0 | 0.00006 | 74 | 0 | 0 | 0 | 29.99 |
| 463 | 49696 | 4 | 0 | 0.00004 | 74 | 0 | 0 | 0 | 30 |
| 464 | 49722 | 1 | 0 | 0.00001 | 74 | 0 | 0 | 0 | 30 |
| 465 | 49387 | 7 | 0 | 0.000071 | 74 | 0 | 0 | 0 | 29.99 |
| 466 | 49799 | 1 | 0 | 0.00001 | 74 | 0 | 0 | 0 | 30 |
| 467 | 49831 | 2 | 0 | 0.00002 | 74 | 0 | 0 | 0 | 30 |
| 468 | 49590 | 190 | 58 | 0.00307 | 73 | 0 | 0 | 0 | 29.67 |
| 469 | 35368 | 8646 | 5857 | 0.204127 | 57 | 11 | 6 | 0.155405 | 20.77 |
| 470 | 49878 | 1 | 0 | 0.00001 | 74 | 0 | 0 | 0 | 50 |
| 471 | 49872 | 1 | 0 | 0.00001 | 74 | 0 | 0 | 0 | 30 |
| 472 | 49801 | 6 | 0 | 0.00006 | 74 | 0 | 0 | 0 | 29.99 |
| 473 | 49761 | 2 | 0 | 0.00002 | 74 | 0 | 0 | 0 | 30 |
| 474 | 49422 | 188 | 17 | 0.002237 | 73 | 0 | 0 | 0 | 29.76 |
| 475 | 49508 | 3 | 0 | 0.00003 | 74 | 0 | 0 | 0 | 30 |
| 476 | 49840 | 1 | 1 | 0.00003 | 74 | 0 | 0 | 0 | 30 |
| 477 | 49839 | 10 | 0 | 0.0001 | 74 | 0 | 0 | 0 | 29.99 |
| 478 | 49873 | 6 | 0 | 0.00006 | 74 | 0 | 0 | 0 | 29.99 |
| 479 | 49876 | 3 | 0 | 0.00003 | 74 | 0 | 0 | 0 | 49.99 |
| 480 | 49839 | 34 | 5 | 0.000441 | 74 | 0 | 0 | 0 | 49.92 |
| 481 | 49877 | 2 | 0 | 0.00002 | 74 | 0 | 0 | 0 | 50 |
| 482 | 49877 | 2 | 0 | 0.00002 | 74 | 0 | 0 | 0 | 50 |
| 483 | 49878 | 1 | 0 | 0.00001 | 74 | 0 | 0 | 0 | 50 |
| 484 | 49875 | 2 | 1 | 0.00004 | 74 | 0 | 0 | 0 | 49.99 |
| 485 | 49877 | 1 | 0 | 0.00001 | 74 | 0 | 0 | 0 | 50 |
| 486 | 49875 | 1 | 0 | 0.00001 | 74 | 0 | 0 | 0 | 30 |
| 487 | 49850 | 3 | 0 | 0.00003 | 74 | 0 | 0 | 0 | 30 |
| 488 | 49522 | 1 | 0 | 0.00001 | 73 | 0 | 0 | 0 | 30 |
| 489 | 49389 | 2 | 0 | 0.00002 | 71 | 0 | 0 | 0 | 30 |
| 490 | 0 | 26 | 49024 | 0.999735 | 0 | 0 | 72 | 1 | 29.97 |
| 491 | 49341 | 15 | 0 | 0.000152 | 74 | 0 | 0 | 0 | 29.98 |
| 492 | 49419 | 1 | 0 | 0.00001 | 74 | 0 | 0 | 0 | 30 |
| 493 | 49522 | 1 | 0 | 0.00001 | 73 | 0 | 0 | 0 | 30 |
| 494 | 49647 | 2 | 0 | 0.00002 | 73 | 0 | 0 | 0 | 30 |
| 495 | 49654 | 2 | 0 | 0.00002 | 73 | 0 | 0 | 0 | 30 |
| 496 | 49857 | 1 | 0 | 0.00001 | 74 | 0 | 0 | 0 | 30 |
| 497 | 49874 | 4 | 1 | 0.00006 | 74 | 0 | 0 | 0 | 49.99 |
| 498 | 49878 | 1 | 0 | 0.00001 | 74 | 0 | 0 | 0 | 50 |
| 499 | 49878 | 1 | 0 | 0.00001 | 74 | 0 | 0 | 0 | 50 |
| 500 | 49878 | 1 | 0 | 0.00001 | 74 | 0 | 0 | 0 | 50 |
| 501 | 49877 | 2 | 0 | 0.00002 | 74 | 0 | 0 | 0 | 50 |
| 502 | 49858 | 17 | 3 | 0.000231 | 74 | 0 | 0 | 0 | 49.96 |
| 503 | 49341 | 377 | 155 | 0.006887 | 73 | 0 | 1 | 0.013514 | 48.77 |
| 504 | 49878 | 1 | 0 | 0.00001 | 74 | 0 | 0 | 0 | 50 |
| 505 | 49873 | 3 | 1 | 0.00005 | 74 | 0 | 0 | 0 | 49.99 |
| 506 | 49874 | 4 | 1 | 0.00006 | 74 | 0 | 0 | 0 | 49.99 |
| 507 | 49875 | 2 | 1 | 0.00004 | 74 | 0 | 0 | 0 | 49.99 |
| 508 | 49875 | 3 | 0 | 0.00003 | 74 | 0 | 0 | 0 | 49.99 |
| 509 | 49851 | 23 | 3 | 0.000291 | 74 | 0 | 0 | 0 | 49.95 |
| 510 | 49832 | 2 | 0 | 0.00002 | 74 | 0 | 0 | 0 | 50 |

**Supplementary Table 1B.** Genotype counts and frequencies for variants in *TMPRSS2*.

| Locus | contAA | contAB | contBB | contFreq | caseAA | caseAB | caseBB | caseFreq | weight |
| --- | --- | --- | --- | --- | --- | --- | --- | --- | --- |
| 1 | 49866 | 1 | 0 | 0.00001 | 74 | 0 | 0 | 0 | 50 |
| 2 | 49870 | 1 | 0 | 0.00001 | 74 | 0 | 0 | 0 | 50 |
| 3 | 49866 | 7 | 0 | 0.00007 | 74 | 0 | 0 | 0 | 49.99 |
| 4 | 49874 | 2 | 0 | 0.00002 | 74 | 0 | 0 | 0 | 50 |
| 5 | 49869 | 10 | 0 | 0.0001 | 74 | 0 | 0 | 0 | 49.98 |
| 6 | 49868 | 11 | 0 | 0.00011 | 74 | 0 | 0 | 0 | 49.98 |
| 7 | 49877 | 2 | 0 | 0.00002 | 74 | 0 | 0 | 0 | 50 |
| 8 | 49877 | 2 | 0 | 0.00002 | 74 | 0 | 0 | 0 | 50 |
| 9 | 49877 | 2 | 0 | 0.00002 | 74 | 0 | 0 | 0 | 50 |
| 10 | 49876 | 3 | 0 | 0.00003 | 74 | 0 | 0 | 0 | 49.99 |
| 11 | 49878 | 1 | 0 | 0.00001 | 74 | 0 | 0 | 0 | 50 |
| 12 | 49877 | 2 | 0 | 0.00002 | 74 | 0 | 0 | 0 | 50 |
| 13 | 49878 | 1 | 0 | 0.00001 | 74 | 0 | 0 | 0 | 50 |
| 14 | 49877 | 2 | 0 | 0.00002 | 74 | 0 | 0 | 0 | 50 |
| 15 | 49878 | 1 | 0 | 0.00001 | 74 | 0 | 0 | 0 | 50 |
| 16 | 49876 | 3 | 0 | 0.00003 | 74 | 0 | 0 | 0 | 49.99 |
| 17 | 49878 | 1 | 0 | 0.00001 | 74 | 0 | 0 | 0 | 50 |
| 18 | 49874 | 5 | 0 | 0.00005 | 74 | 0 | 0 | 0 | 49.99 |
| 19 | 49856 | 23 | 0 | 0.000231 | 74 | 0 | 0 | 0 | 49.96 |
| 20 | 49877 | 2 | 0 | 0.00002 | 74 | 0 | 0 | 0 | 50 |
| 21 | 49868 | 11 | 0 | 0.00011 | 74 | 0 | 0 | 0 | 49.98 |
| 22 | 49878 | 1 | 0 | 0.00001 | 74 | 0 | 0 | 0 | 50 |
| 23 | 49876 | 3 | 0 | 0.00003 | 74 | 0 | 0 | 0 | 30 |
| 24 | 49878 | 1 | 0 | 0.00001 | 74 | 0 | 0 | 0 | 30 |
| 25 | 49878 | 1 | 0 | 0.00001 | 74 | 0 | 0 | 0 | 30 |
| 26 | 0 | 24 | 45793 | 0.999738 | 0 | 0 | 71 | 1 | 29.97 |
| 27 | 49878 | 1 | 0 | 0.00001 | 74 | 0 | 0 | 0 | 30 |
| 28 | 49859 | 20 | 0 | 0.0002 | 74 | 0 | 0 | 0 | 29.98 |
| 29 | 49815 | 62 | 2 | 0.000662 | 74 | 0 | 0 | 0 | 29.93 |
| 30 | 49877 | 2 | 0 | 0.00002 | 74 | 0 | 0 | 0 | 30 |
| 31 | 49878 | 1 | 0 | 0.00001 | 74 | 0 | 0 | 0 | 30 |
| 32 | 49876 | 3 | 0 | 0.00003 | 74 | 0 | 0 | 0 | 30 |
| 33 | 49878 | 1 | 0 | 0.00001 | 74 | 0 | 0 | 0 | 30 |
| 34 | 49875 | 3 | 0 | 0.00003 | 74 | 0 | 0 | 0 | 30 |
| 35 | 49874 | 4 | 0 | 0.00004 | 74 | 0 | 0 | 0 | 30 |
| 36 | 38028 | 11037 | 812 | 0.126922 | 62 | 12 | 0 | 0.081081 | 18.04 |
| 37 | 49842 | 8 | 0 | 0.00008 | 74 | 0 | 0 | 0 | 29.99 |
| 38 | 49830 | 1 | 0 | 0.00001 | 74 | 0 | 0 | 0 | 30 |
| 39 | 49816 | 8 | 0 | 0.00008 | 74 | 0 | 0 | 0 | 29.99 |
| 40 | 49761 | 1 | 0 | 0.00001 | 74 | 0 | 0 | 0 | 30 |
| 41 | 49878 | 1 | 0 | 0.00001 | 74 | 0 | 0 | 0 | 30 |
| 42 | 49877 | 2 | 0 | 0.00002 | 74 | 0 | 0 | 0 | 30 |
| 43 | 49874 | 5 | 0 | 0.00005 | 74 | 0 | 0 | 0 | 29.99 |
| 44 | 49681 | 198 | 0 | 0.001985 | 74 | 0 | 0 | 0 | 29.79 |
| 45 | 49877 | 2 | 0 | 0.00002 | 74 | 0 | 0 | 0 | 30 |
| 46 | 49878 | 1 | 0 | 0.00001 | 74 | 0 | 0 | 0 | 30 |
| 47 | 49878 | 1 | 0 | 0.00001 | 74 | 0 | 0 | 0 | 30 |
| 48 | 49878 | 1 | 0 | 0.00001 | 74 | 0 | 0 | 0 | 30 |
| 49 | 49779 | 100 | 0 | 0.001002 | 74 | 0 | 0 | 0 | 29.89 |
| 50 | 49877 | 2 | 0 | 0.00002 | 74 | 0 | 0 | 0 | 30 |
| 51 | 49878 | 1 | 0 | 0.00001 | 74 | 0 | 0 | 0 | 30 |
| 52 | 49878 | 1 | 0 | 0.00001 | 74 | 0 | 0 | 0 | 50 |
| 53 | 49164 | 713 | 2 | 0.007187 | 74 | 0 | 0 | 0 | 48.72 |
| 54 | 49877 | 2 | 0 | 0.00002 | 74 | 0 | 0 | 0 | 50 |
| 55 | 49877 | 2 | 0 | 0.00002 | 74 | 0 | 0 | 0 | 50 |
| 56 | 49877 | 2 | 0 | 0.00002 | 74 | 0 | 0 | 0 | 50 |
| 57 | 49876 | 3 | 0 | 0.00003 | 74 | 0 | 0 | 0 | 49.99 |
| 58 | 49878 | 1 | 0 | 0.00001 | 74 | 0 | 0 | 0 | 50 |
| 59 | 49875 | 4 | 0 | 0.00004 | 74 | 0 | 0 | 0 | 49.99 |
| 60 | 49878 | 1 | 0 | 0.00001 | 74 | 0 | 0 | 0 | 50 |
| 61 | 49867 | 12 | 0 | 0.00012 | 74 | 0 | 0 | 0 | 49.98 |
| 62 | 49872 | 7 | 0 | 0.00007 | 74 | 0 | 0 | 0 | 49.99 |
| 63 | 49878 | 1 | 0 | 0.00001 | 74 | 0 | 0 | 0 | 50 |
| 64 | 49878 | 1 | 0 | 0.00001 | 74 | 0 | 0 | 0 | 30 |
| 65 | 49878 | 1 | 0 | 0.00001 | 74 | 0 | 0 | 0 | 30 |
| 66 | 49485 | 391 | 3 | 0.00398 | 74 | 0 | 0 | 0 | 29.57 |
| 67 | 49878 | 1 | 0 | 0.00001 | 74 | 0 | 0 | 0 | 30 |
| 68 | 49613 | 265 | 1 | 0.002676 | 74 | 0 | 0 | 0 | 29.71 |
| 69 | 49878 | 1 | 0 | 0.00001 | 74 | 0 | 0 | 0 | 30 |
| 70 | 49853 | 25 | 0 | 0.000251 | 74 | 0 | 0 | 0 | 29.97 |
| 71 | 49875 | 2 | 0 | 0.00002 | 74 | 0 | 0 | 0 | 30 |
| 72 | 49845 | 2 | 0 | 0.00002 | 74 | 0 | 0 | 0 | 30 |
| 73 | 49845 | 8 | 0 | 0.00008 | 74 | 0 | 0 | 0 | 29.99 |
| 74 | 49845 | 1 | 0 | 0.00001 | 74 | 0 | 0 | 0 | 30 |
| 75 | 49872 | 2 | 0 | 0.00002 | 74 | 0 | 0 | 0 | 30 |
| 76 | 49872 | 1 | 0 | 0.00001 | 74 | 0 | 0 | 0 | 30 |
| 77 | 49864 | 13 | 0 | 0.00013 | 74 | 0 | 0 | 0 | 29.99 |
| 78 | 49878 | 1 | 0 | 0.00001 | 74 | 0 | 0 | 0 | 30 |
| 79 | 49878 | 1 | 0 | 0.00001 | 74 | 0 | 0 | 0 | 30 |
| 80 | 49877 | 2 | 0 | 0.00002 | 74 | 0 | 0 | 0 | 30 |
| 81 | 49870 | 9 | 0 | 0.00009 | 74 | 0 | 0 | 0 | 29.99 |
| 82 | 49878 | 1 | 0 | 0.00001 | 74 | 0 | 0 | 0 | 30 |
| 83 | 49878 | 1 | 0 | 0.00001 | 74 | 0 | 0 | 0 | 30 |
| 84 | 49878 | 1 | 0 | 0.00001 | 74 | 0 | 0 | 0 | 30 |
| 85 | 49878 | 1 | 0 | 0.00001 | 74 | 0 | 0 | 0 | 30 |
| 86 | 49878 | 1 | 0 | 0.00001 | 74 | 0 | 0 | 0 | 30 |
| 87 | 49874 | 5 | 0 | 0.00005 | 74 | 0 | 0 | 0 | 29.99 |
| 88 | 49878 | 1 | 0 | 0.00001 | 74 | 0 | 0 | 0 | 30 |
| 89 | 49878 | 1 | 0 | 0.00001 | 74 | 0 | 0 | 0 | 199.99 |
| 90 | 49877 | 2 | 0 | 0.00002 | 74 | 0 | 0 | 0 | 50 |
| 91 | 49876 | 3 | 0 | 0.00003 | 74 | 0 | 0 | 0 | 49.99 |
| 92 | 49878 | 1 | 0 | 0.00001 | 74 | 0 | 0 | 0 | 50 |
| 93 | 49878 | 1 | 0 | 0.00001 | 74 | 0 | 0 | 0 | 50 |
| 94 | 49876 | 3 | 0 | 0.00003 | 74 | 0 | 0 | 0 | 49.99 |
| 95 | 49877 | 2 | 0 | 0.00002 | 74 | 0 | 0 | 0 | 50 |
| 96 | 49878 | 1 | 0 | 0.00001 | 74 | 0 | 0 | 0 | 50 |
| 97 | 49718 | 160 | 1 | 0.001624 | 74 | 0 | 0 | 0 | 49.71 |
| 98 | 49878 | 1 | 0 | 0.00001 | 74 | 0 | 0 | 0 | 50 |
| 99 | 49878 | 1 | 0 | 0.00001 | 74 | 0 | 0 | 0 | 50 |
| 100 | 49878 | 1 | 0 | 0.00001 | 74 | 0 | 0 | 0 | 50 |
| 101 | 49878 | 1 | 0 | 0.00001 | 74 | 0 | 0 | 0 | 50 |
| 102 | 49878 | 1 | 0 | 0.00001 | 74 | 0 | 0 | 0 | 50 |
| 103 | 49878 | 1 | 0 | 0.00001 | 74 | 0 | 0 | 0 | 50 |
| 104 | 49878 | 1 | 0 | 0.00001 | 74 | 0 | 0 | 0 | 50 |
| 105 | 49878 | 1 | 0 | 0.00001 | 74 | 0 | 0 | 0 | 50 |
| 106 | 49878 | 1 | 0 | 0.00001 | 74 | 0 | 0 | 0 | 50 |
| 107 | 49878 | 1 | 0 | 0.00001 | 74 | 0 | 0 | 0 | 50 |
| 108 | 49878 | 1 | 0 | 0.00001 | 74 | 0 | 0 | 0 | 50 |
| 109 | 49877 | 2 | 0 | 0.00002 | 74 | 0 | 0 | 0 | 50 |
| 110 | 49878 | 1 | 0 | 0.00001 | 74 | 0 | 0 | 0 | 50 |
| 111 | 49877 | 2 | 0 | 0.00002 | 74 | 0 | 0 | 0 | 50 |
| 112 | 49878 | 1 | 0 | 0.00001 | 74 | 0 | 0 | 0 | 30 |
| 113 | 49878 | 1 | 0 | 0.00001 | 74 | 0 | 0 | 0 | 30 |
| 114 | 49857 | 22 | 0 | 0.000221 | 73 | 1 | 0 | 0.006757 | 29.98 |
| 115 | 49878 | 1 | 0 | 0.00001 | 74 | 0 | 0 | 0 | 30 |
| 116 | 49878 | 1 | 0 | 0.00001 | 74 | 0 | 0 | 0 | 30 |
| 117 | 49872 | 7 | 0 | 0.00007 | 74 | 0 | 0 | 0 | 29.99 |
| 118 | 49878 | 1 | 0 | 0.00001 | 74 | 0 | 0 | 0 | 30 |
| 119 | 49878 | 1 | 0 | 0.00001 | 74 | 0 | 0 | 0 | 30 |
| 120 | 49878 | 1 | 0 | 0.00001 | 74 | 0 | 0 | 0 | 30 |
| 121 | 49878 | 1 | 0 | 0.00001 | 74 | 0 | 0 | 0 | 30 |
| 122 | 49878 | 1 | 0 | 0.00001 | 74 | 0 | 0 | 0 | 30 |
| 123 | 49878 | 1 | 0 | 0.00001 | 74 | 0 | 0 | 0 | 30 |
| 124 | 49843 | 1 | 0 | 0.00001 | 74 | 0 | 0 | 0 | 30 |
| 125 | 49875 | 1 | 0 | 0.00001 | 74 | 0 | 0 | 0 | 30 |
| 126 | 49872 | 4 | 0 | 0.00004 | 74 | 0 | 0 | 0 | 30 |
| 127 | 49782 | 94 | 3 | 0.001002 | 73 | 1 | 0 | 0.006757 | 29.89 |
| 128 | 49878 | 1 | 0 | 0.00001 | 74 | 0 | 0 | 0 | 30 |
| 129 | 49875 | 4 | 0 | 0.00004 | 74 | 0 | 0 | 0 | 30 |
| 130 | 49877 | 2 | 0 | 0.00002 | 74 | 0 | 0 | 0 | 30 |
| 131 | 49878 | 1 | 0 | 0.00001 | 74 | 0 | 0 | 0 | 50 |
| 132 | 49873 | 6 | 0 | 0.00006 | 74 | 0 | 0 | 0 | 49.99 |
| 133 | 47048 | 2782 | 49 | 0.02887 | 72 | 2 | 0 | 0.013514 | 44.96 |
| 134 | 49877 | 1 | 0 | 0.00001 | 74 | 0 | 0 | 0 | 299.99 |
| 135 | 49878 | 1 | 0 | 0.00001 | 74 | 0 | 0 | 0 | 50 |
| 136 | 49872 | 7 | 0 | 0.00007 | 74 | 0 | 0 | 0 | 49.99 |
| 137 | 49877 | 2 | 0 | 0.00002 | 74 | 0 | 0 | 0 | 50 |
| 138 | 49878 | 1 | 0 | 0.00001 | 74 | 0 | 0 | 0 | 50 |
| 139 | 49872 | 7 | 0 | 0.00007 | 74 | 0 | 0 | 0 | 49.99 |
| 140 | 49876 | 3 | 0 | 0.00003 | 74 | 0 | 0 | 0 | 49.99 |
| 141 | 49868 | 2 | 0 | 0.00002 | 74 | 0 | 0 | 0 | 50 |
| 142 | 49878 | 1 | 0 | 0.00001 | 74 | 0 | 0 | 0 | 50 |
| 143 | 49878 | 1 | 0 | 0.00001 | 74 | 0 | 0 | 0 | 50 |
| 144 | 49878 | 1 | 0 | 0.00001 | 74 | 0 | 0 | 0 | 199.99 |
| 145 | 49877 | 2 | 0 | 0.00002 | 74 | 0 | 0 | 0 | 50 |
| 146 | 49878 | 1 | 0 | 0.00001 | 74 | 0 | 0 | 0 | 50 |
| 147 | 49878 | 1 | 0 | 0.00001 | 74 | 0 | 0 | 0 | 30 |
| 148 | 49878 | 1 | 0 | 0.00001 | 74 | 0 | 0 | 0 | 30 |
| 149 | 49855 | 24 | 0 | 0.000241 | 74 | 0 | 0 | 0 | 29.97 |
| 150 | 49842 | 37 | 0 | 0.000371 | 74 | 0 | 0 | 0 | 29.96 |
| 151 | 49878 | 1 | 0 | 0.00001 | 74 | 0 | 0 | 0 | 30 |
| 152 | 49823 | 20 | 0 | 0.000201 | 73 | 1 | 0 | 0.006757 | 29.98 |
| 153 | 49874 | 5 | 0 | 0.00005 | 74 | 0 | 0 | 0 | 29.99 |
| 154 | 49875 | 1 | 0 | 0.00001 | 74 | 0 | 0 | 0 | 30 |
| 155 | 49876 | 1 | 0 | 0.00001 | 74 | 0 | 0 | 0 | 30 |
| 156 | 49832 | 9 | 0 | 0.00009 | 74 | 0 | 0 | 0 | 29.99 |
| 157 | 49821 | 2 | 0 | 0.00002 | 74 | 0 | 0 | 0 | 30 |
| 158 | 49755 | 20 | 0 | 0.000201 | 74 | 0 | 0 | 0 | 29.98 |
| 159 | 49875 | 2 | 0 | 0.00002 | 74 | 0 | 0 | 0 | 30 |
| 160 | 49875 | 2 | 0 | 0.00002 | 74 | 0 | 0 | 0 | 30 |
| 161 | 49876 | 1 | 0 | 0.00001 | 74 | 0 | 0 | 0 | 30 |
| 162 | 49869 | 8 | 1 | 0.0001 | 74 | 0 | 0 | 0 | 29.99 |
| 163 | 49878 | 1 | 0 | 0.00001 | 74 | 0 | 0 | 0 | 30 |
| 164 | 49873 | 6 | 0 | 0.00006 | 74 | 0 | 0 | 0 | 29.99 |
| 165 | 49878 | 1 | 0 | 0.00001 | 74 | 0 | 0 | 0 | 30 |
| 166 | 49876 | 3 | 0 | 0.00003 | 74 | 0 | 0 | 0 | 30 |
| 167 | 49874 | 5 | 0 | 0.00005 | 74 | 0 | 0 | 0 | 29.99 |
| 168 | 49878 | 1 | 0 | 0.00001 | 74 | 0 | 0 | 0 | 50 |
| 169 | 49878 | 1 | 0 | 0.00001 | 74 | 0 | 0 | 0 | 50 |
| 170 | 49877 | 2 | 0 | 0.00002 | 74 | 0 | 0 | 0 | 50 |
| 171 | 49876 | 3 | 0 | 0.00003 | 74 | 0 | 0 | 0 | 49.99 |
| 172 | 49876 | 3 | 0 | 0.00003 | 74 | 0 | 0 | 0 | 49.99 |
| 173 | 49878 | 1 | 0 | 0.00001 | 74 | 0 | 0 | 0 | 50 |
| 174 | 49878 | 1 | 0 | 0.00001 | 74 | 0 | 0 | 0 | 50 |
| 175 | 49876 | 3 | 0 | 0.00003 | 74 | 0 | 0 | 0 | 49.99 |
| 176 | 49878 | 1 | 0 | 0.00001 | 74 | 0 | 0 | 0 | 50 |
| 177 | 49878 | 1 | 0 | 0.00001 | 74 | 0 | 0 | 0 | 50 |
| 178 | 49878 | 1 | 0 | 0.00001 | 74 | 0 | 0 | 0 | 50 |
| 179 | 49878 | 1 | 0 | 0.00001 | 74 | 0 | 0 | 0 | 50 |
| 180 | 49878 | 1 | 0 | 0.00001 | 74 | 0 | 0 | 0 | 50 |
| 181 | 49878 | 1 | 0 | 0.00001 | 74 | 0 | 0 | 0 | 50 |
| 182 | 49878 | 1 | 0 | 0.00001 | 74 | 0 | 0 | 0 | 50 |
| 183 | 49878 | 1 | 0 | 0.00001 | 74 | 0 | 0 | 0 | 50 |
| 184 | 49860 | 19 | 0 | 0.00019 | 74 | 0 | 0 | 0 | 49.97 |
| 185 | 49877 | 2 | 0 | 0.00002 | 74 | 0 | 0 | 0 | 50 |
| 186 | 49877 | 2 | 0 | 0.00002 | 74 | 0 | 0 | 0 | 50 |
| 187 | 49877 | 2 | 0 | 0.00002 | 74 | 0 | 0 | 0 | 50 |
| 188 | 49861 | 7 | 0 | 0.00007 | 74 | 0 | 0 | 0 | 99.97 |
| 189 | 49873 | 6 | 0 | 0.00006 | 74 | 0 | 0 | 0 | 49.99 |
| 190 | 49878 | 1 | 0 | 0.00001 | 74 | 0 | 0 | 0 | 50 |
| 191 | 49876 | 1 | 0 | 0.00001 | 74 | 0 | 0 | 0 | 50 |
| 192 | 49878 | 1 | 0 | 0.00001 | 74 | 0 | 0 | 0 | 50 |
| 193 | 49875 | 4 | 0 | 0.00004 | 74 | 0 | 0 | 0 | 49.99 |
| 194 | 49870 | 9 | 0 | 0.00009 | 74 | 0 | 0 | 0 | 49.98 |
| 195 | 49877 | 2 | 0 | 0.00002 | 74 | 0 | 0 | 0 | 50 |
| 196 | 49878 | 1 | 0 | 0.00001 | 74 | 0 | 0 | 0 | 50 |
| 197 | 49877 | 2 | 0 | 0.00002 | 74 | 0 | 0 | 0 | 50 |
| 198 | 49878 | 1 | 0 | 0.00001 | 74 | 0 | 0 | 0 | 30 |
| 199 | 49878 | 1 | 0 | 0.00001 | 74 | 0 | 0 | 0 | 30 |
| 200 | 49749 | 130 | 0 | 0.001303 | 74 | 0 | 0 | 0 | 29.86 |
| 201 | 49843 | 36 | 0 | 0.000361 | 74 | 0 | 0 | 0 | 29.96 |
| 202 | 49870 | 9 | 0 | 0.00009 | 74 | 0 | 0 | 0 | 29.99 |
| 203 | 49878 | 1 | 0 | 0.00001 | 74 | 0 | 0 | 0 | 30 |
| 204 | 48473 | 1337 | 64 | 0.014687 | 71 | 3 | 0 | 0.02027 | 28.44 |
| 205 | 16267 | 24053 | 9559 | 0.432757 | 26 | 34 | 14 | 0.418919 | 3.49 |
| 206 | 49877 | 1 | 0 | 0.00001 | 74 | 0 | 0 | 0 | 30 |
| 207 | 49873 | 4 | 0 | 0.00004 | 74 | 0 | 0 | 0 | 30 |
| 208 | 49873 | 2 | 0 | 0.00002 | 74 | 0 | 0 | 0 | 30 |
| 209 | 49874 | 1 | 0 | 0.00001 | 74 | 0 | 0 | 0 | 30 |
| 210 | 49875 | 2 | 0 | 0.00002 | 74 | 0 | 0 | 0 | 30 |
| 211 | 49874 | 3 | 0 | 0.00003 | 74 | 0 | 0 | 0 | 30 |
| 212 | 49826 | 6 | 0 | 0.00006 | 74 | 0 | 0 | 0 | 29.99 |
| 213 | 46375 | 3431 | 69 | 0.035779 | 69 | 5 | 0 | 0.033784 | 26.27 |
| 214 | 49878 | 1 | 0 | 0.00001 | 74 | 0 | 0 | 0 | 30 |
| 215 | 49878 | 1 | 0 | 0.00001 | 74 | 0 | 0 | 0 | 30 |
| 216 | 49878 | 1 | 0 | 0.00001 | 74 | 0 | 0 | 0 | 30 |
| 217 | 49878 | 1 | 0 | 0.00001 | 74 | 0 | 0 | 0 | 30 |
| 218 | 49860 | 19 | 0 | 0.00019 | 74 | 0 | 0 | 0 | 29.98 |
| 219 | 49443 | 422 | 14 | 0.004511 | 72 | 2 | 0 | 0.013514 | 29.51 |
| 220 | 49426 | 438 | 15 | 0.004691 | 72 | 2 | 0 | 0.013514 | 29.49 |
| 221 | 48377 | 1484 | 18 | 0.015237 | 73 | 1 | 0 | 0.006757 | 28.38 |
| 222 | 49872 | 7 | 0 | 0.00007 | 74 | 0 | 0 | 0 | 29.99 |
| 223 | 49872 | 1 | 0 | 0.00001 | 74 | 0 | 0 | 0 | 30 |
| 224 | 49876 | 3 | 0 | 0.00003 | 74 | 0 | 0 | 0 | 30 |
| 225 | 49878 | 1 | 0 | 0.00001 | 74 | 0 | 0 | 0 | 30 |
| 226 | 49865 | 14 | 0 | 0.00014 | 74 | 0 | 0 | 0 | 29.98 |
| 227 | 49846 | 33 | 0 | 0.000331 | 74 | 0 | 0 | 0 | 29.96 |
| 228 | 49878 | 1 | 0 | 0.00001 | 74 | 0 | 0 | 0 | 30 |
| 229 | 49878 | 1 | 0 | 0.00001 | 74 | 0 | 0 | 0 | 30 |
| 230 | 49878 | 1 | 0 | 0.00001 | 74 | 0 | 0 | 0 | 50 |
| 231 | 49864 | 15 | 0 | 0.00015 | 74 | 0 | 0 | 0 | 49.97 |
| 232 | 49875 | 4 | 0 | 0.00004 | 74 | 0 | 0 | 0 | 49.99 |
| 233 | 49871 | 8 | 0 | 0.00008 | 74 | 0 | 0 | 0 | 49.99 |
| 234 | 49878 | 1 | 0 | 0.00001 | 74 | 0 | 0 | 0 | 50 |
| 235 | 49878 | 1 | 0 | 0.00001 | 74 | 0 | 0 | 0 | 50 |
| 236 | 49878 | 1 | 0 | 0.00001 | 74 | 0 | 0 | 0 | 50 |
| 237 | 49878 | 1 | 0 | 0.00001 | 74 | 0 | 0 | 0 | 50 |
| 238 | 49878 | 1 | 0 | 0.00001 | 74 | 0 | 0 | 0 | 50 |
| 239 | 49878 | 1 | 0 | 0.00001 | 74 | 0 | 0 | 0 | 50 |
| 240 | 49871 | 8 | 0 | 0.00008 | 74 | 0 | 0 | 0 | 49.99 |
| 241 | 49878 | 1 | 0 | 0.00001 | 74 | 0 | 0 | 0 | 50 |
| 242 | 49878 | 1 | 0 | 0.00001 | 74 | 0 | 0 | 0 | 50 |
| 243 | 49874 | 5 | 0 | 0.00005 | 74 | 0 | 0 | 0 | 49.99 |
| 244 | 49877 | 2 | 0 | 0.00002 | 74 | 0 | 0 | 0 | 50 |
| 245 | 49878 | 1 | 0 | 0.00001 | 74 | 0 | 0 | 0 | 50 |
| 246 | 49878 | 1 | 0 | 0.00001 | 74 | 0 | 0 | 0 | 50 |
| 247 | 49873 | 6 | 0 | 0.00006 | 74 | 0 | 0 | 0 | 49.99 |
| 248 | 49875 | 4 | 0 | 0.00004 | 74 | 0 | 0 | 0 | 49.99 |
| 249 | 49870 | 9 | 0 | 0.00009 | 74 | 0 | 0 | 0 | 49.98 |
| 250 | 49877 | 2 | 0 | 0.00002 | 74 | 0 | 0 | 0 | 50 |
| 251 | 49860 | 2 | 0 | 0.00002 | 74 | 0 | 0 | 0 | 99.99 |
| 252 | 49878 | 1 | 0 | 0.00001 | 74 | 0 | 0 | 0 | 50 |
| 253 | 49878 | 1 | 0 | 0.00001 | 74 | 0 | 0 | 0 | 50 |
| 254 | 49877 | 2 | 0 | 0.00002 | 74 | 0 | 0 | 0 | 50 |
| 255 | 49878 | 1 | 0 | 0.00001 | 74 | 0 | 0 | 0 | 50 |
| 256 | 49870 | 9 | 0 | 0.00009 | 74 | 0 | 0 | 0 | 49.98 |
| 257 | 49877 | 2 | 0 | 0.00002 | 74 | 0 | 0 | 0 | 50 |
| 258 | 49862 | 17 | 0 | 0.00017 | 74 | 0 | 0 | 0 | 49.97 |
| 259 | 49877 | 2 | 0 | 0.00002 | 74 | 0 | 0 | 0 | 50 |
| 260 | 49867 | 12 | 0 | 0.00012 | 74 | 0 | 0 | 0 | 49.98 |
| 261 | 49876 | 1 | 0 | 0.00001 | 74 | 0 | 0 | 0 | 50 |
| 262 | 49869 | 10 | 0 | 0.0001 | 74 | 0 | 0 | 0 | 49.98 |
| 263 | 29807 | 17442 | 2630 | 0.227571 | 51 | 22 | 1 | 0.162162 | 18.37 |
| 264 | 49867 | 1 | 0 | 0.00001 | 74 | 0 | 0 | 0 | 299.99 |
| 265 | 49878 | 1 | 0 | 0.00001 | 74 | 0 | 0 | 0 | 50 |
| 266 | 14707 | 24543 | 10629 | 0.459121 | 24 | 36 | 14 | 0.432432 | 5.3 |
| 267 | 49878 | 1 | 0 | 0.00001 | 74 | 0 | 0 | 0 | 50 |
| 268 | 49878 | 1 | 0 | 0.00001 | 74 | 0 | 0 | 0 | 50 |
| 269 | 49878 | 1 | 0 | 0.00001 | 74 | 0 | 0 | 0 | 50 |
| 270 | 49878 | 1 | 0 | 0.00001 | 74 | 0 | 0 | 0 | 50 |
| 271 | 49878 | 1 | 0 | 0.00001 | 74 | 0 | 0 | 0 | 30 |
| 272 | 49875 | 4 | 0 | 0.00004 | 74 | 0 | 0 | 0 | 30 |
| 273 | 49878 | 1 | 0 | 0.00001 | 74 | 0 | 0 | 0 | 30 |
| 274 | 49872 | 7 | 0 | 0.00007 | 74 | 0 | 0 | 0 | 29.99 |
| 275 | 49878 | 1 | 0 | 0.00001 | 74 | 0 | 0 | 0 | 30 |
| 276 | 49878 | 1 | 0 | 0.00001 | 74 | 0 | 0 | 0 | 30 |
| 277 | 49823 | 56 | 0 | 0.000561 | 74 | 0 | 0 | 0 | 29.94 |
| 278 | 49878 | 1 | 0 | 0.00001 | 74 | 0 | 0 | 0 | 30 |
| 279 | 49876 | 1 | 0 | 0.00001 | 74 | 0 | 0 | 0 | 30 |
| 280 | 49870 | 7 | 0 | 0.00007 | 74 | 0 | 0 | 0 | 29.99 |
| 281 | 49874 | 3 | 0 | 0.00003 | 74 | 0 | 0 | 0 | 30 |
| 282 | 49871 | 6 | 0 | 0.00006 | 74 | 0 | 0 | 0 | 29.99 |
| 283 | 49862 | 1 | 0 | 0.00001 | 74 | 0 | 0 | 0 | 30 |
| 284 | 49856 | 3 | 0 | 0.00003 | 74 | 0 | 0 | 0 | 30 |
| 285 | 49846 | 14 | 0 | 0.00014 | 74 | 0 | 0 | 0 | 29.98 |
| 286 | 49836 | 4 | 0 | 0.00004 | 74 | 0 | 0 | 0 | 30 |
| 287 | 49861 | 1 | 0 | 0.00001 | 74 | 0 | 0 | 0 | 30 |
| 288 | 49862 | 1 | 0 | 0.00001 | 74 | 0 | 0 | 0 | 30 |
| 289 | 49866 | 1 | 0 | 0.00001 | 74 | 0 | 0 | 0 | 30 |
| 290 | 49878 | 1 | 0 | 0.00001 | 74 | 0 | 0 | 0 | 30 |
| 291 | 49878 | 1 | 0 | 0.00001 | 74 | 0 | 0 | 0 | 30 |
| 292 | 49309 | 564 | 4 | 0.005734 | 74 | 0 | 0 | 0 | 29.39 |
| 293 | 49878 | 1 | 0 | 0.00001 | 74 | 0 | 0 | 0 | 30 |
| 294 | 49876 | 3 | 0 | 0.00003 | 74 | 0 | 0 | 0 | 30 |
| 295 | 49864 | 15 | 0 | 0.00015 | 72 | 2 | 0 | 0.013514 | 29.98 |
| 296 | 49878 | 1 | 0 | 0.00001 | 74 | 0 | 0 | 0 | 50 |
| 297 | 49878 | 1 | 0 | 0.00001 | 74 | 0 | 0 | 0 | 199.99 |
| 298 | 49877 | 2 | 0 | 0.00002 | 74 | 0 | 0 | 0 | 50 |
| 299 | 49878 | 1 | 0 | 0.00001 | 74 | 0 | 0 | 0 | 50 |
| 300 | 49878 | 1 | 0 | 0.00001 | 74 | 0 | 0 | 0 | 50 |
| 301 | 49878 | 1 | 0 | 0.00001 | 74 | 0 | 0 | 0 | 50 |
| 302 | 47639 | 2021 | 139 | 0.023083 | 67 | 6 | 1 | 0.054054 | 27.56 |
| 303 | 49878 | 1 | 0 | 0.00001 | 74 | 0 | 0 | 0 | 30 |
| 304 | 49875 | 3 | 0 | 0.00003 | 74 | 0 | 0 | 0 | 30 |
| 305 | 47116 | 2716 | 46 | 0.028149 | 70 | 4 | 0 | 0.027027 | 27.05 |
| 306 | 49876 | 1 | 0 | 0.00001 | 74 | 0 | 0 | 0 | 30 |
| 307 | 49869 | 1 | 0 | 0.00001 | 74 | 0 | 0 | 0 | 30 |
| 308 | 49838 | 27 | 0 | 0.000271 | 74 | 0 | 0 | 0 | 29.97 |
| 309 | 49807 | 1 | 0 | 0.00001 | 74 | 0 | 0 | 0 | 30 |
| 310 | 49708 | 46 | 0 | 0.000462 | 74 | 0 | 0 | 0 | 29.95 |
| 311 | 49632 | 1 | 0 | 0.00001 | 74 | 0 | 0 | 0 | 30 |
| 312 | 49553 | 2 | 0 | 0.00002 | 74 | 0 | 0 | 0 | 30 |
| 313 | 29186 | 16538 | 3847 | 0.244417 | 39 | 26 | 9 | 0.297297 | 10.05 |
| 314 | 49842 | 2 | 0 | 0.00002 | 74 | 0 | 0 | 0 | 30 |
| 315 | 49857 | 1 | 0 | 0.00001 | 74 | 0 | 0 | 0 | 30 |
| 316 | 49863 | 1 | 0 | 0.00001 | 74 | 0 | 0 | 0 | 30 |
| 317 | 49876 | 2 | 0 | 0.00002 | 74 | 0 | 0 | 0 | 30 |
| 318 | 49876 | 3 | 0 | 0.00003 | 74 | 0 | 0 | 0 | 49.99 |
| 319 | 49877 | 1 | 0 | 0.00001 | 74 | 0 | 0 | 0 | 299.99 |
| 320 | 49877 | 2 | 0 | 0.00002 | 74 | 0 | 0 | 0 | 50 |
| 321 | 49877 | 2 | 0 | 0.00002 | 74 | 0 | 0 | 0 | 50 |
| 322 | 49638 | 241 | 0 | 0.002416 | 73 | 1 | 0 | 0.006757 | 49.57 |
| 323 | 49857 | 22 | 0 | 0.000221 | 74 | 0 | 0 | 0 | 49.96 |
| 324 | 49876 | 3 | 0 | 0.00003 | 74 | 0 | 0 | 0 | 49.99 |
| 325 | 49877 | 1 | 0 | 0.00001 | 74 | 0 | 0 | 0 | 50 |
| 326 | 49878 | 1 | 0 | 0.00001 | 74 | 0 | 0 | 0 | 50 |
| 327 | 49878 | 1 | 0 | 0.00001 | 74 | 0 | 0 | 0 | 50 |
| 328 | 49878 | 1 | 0 | 0.00001 | 74 | 0 | 0 | 0 | 50 |
| 329 | 49876 | 3 | 0 | 0.00003 | 74 | 0 | 0 | 0 | 199.98 |
| 330 | 49878 | 1 | 0 | 0.00001 | 74 | 0 | 0 | 0 | 30 |
| 331 | 49877 | 1 | 0 | 0.00001 | 74 | 0 | 0 | 0 | 30 |
| 332 | 49875 | 1 | 0 | 0.00001 | 74 | 0 | 0 | 0 | 30 |
| 333 | 49874 | 1 | 0 | 0.00001 | 74 | 0 | 0 | 0 | 30 |
| 334 | 49847 | 24 | 0 | 0.000241 | 74 | 0 | 0 | 0 | 29.97 |
| 335 | 49805 | 57 | 0 | 0.000572 | 74 | 0 | 0 | 0 | 29.94 |
| 336 | 49831 | 1 | 0 | 0.00001 | 74 | 0 | 0 | 0 | 30 |
| 337 | 49832 | 1 | 0 | 0.00001 | 74 | 0 | 0 | 0 | 30 |
| 338 | 49825 | 5 | 0 | 0.00005 | 74 | 0 | 0 | 0 | 29.99 |
| 339 | 49819 | 6 | 0 | 0.00006 | 74 | 0 | 0 | 0 | 29.99 |
| 340 | 49824 | 1 | 0 | 0.00001 | 74 | 0 | 0 | 0 | 30 |
| 341 | 49809 | 6 | 0 | 0.00006 | 74 | 0 | 0 | 0 | 29.99 |
| 342 | 49816 | 1 | 0 | 0.00001 | 74 | 0 | 0 | 0 | 30 |
| 343 | 49878 | 1 | 0 | 0.00001 | 74 | 0 | 0 | 0 | 30 |
| 344 | 49878 | 1 | 0 | 0.00001 | 74 | 0 | 0 | 0 | 30 |
| 345 | 44995 | 4768 | 116 | 0.050121 | 70 | 4 | 0 | 0.027027 | 24.86 |
| 346 | 49866 | 13 | 0 | 0.00013 | 74 | 0 | 0 | 0 | 29.99 |
| 347 | 49869 | 10 | 0 | 0.0001 | 74 | 0 | 0 | 0 | 29.99 |
| 348 | 13592 | 24519 | 11768 | 0.481716 | 21 | 34 | 19 | 0.486486 | 3.04 |
| 349 | 49875 | 4 | 0 | 0.00004 | 74 | 0 | 0 | 0 | 30 |
| 350 | 49758 | 120 | 1 | 0.001223 | 74 | 0 | 0 | 0 | 29.87 |
| 351 | 49878 | 1 | 0 | 0.00001 | 74 | 0 | 0 | 0 | 30 |
| 352 | 49859 | 20 | 0 | 0.0002 | 74 | 0 | 0 | 0 | 29.98 |
| 353 | 49878 | 1 | 0 | 0.00001 | 74 | 0 | 0 | 0 | 30 |
| 354 | 49873 | 6 | 0 | 0.00006 | 74 | 0 | 0 | 0 | 49.99 |
| 355 | 49878 | 1 | 0 | 0.00001 | 74 | 0 | 0 | 0 | 199.99 |
| 356 | 49878 | 1 | 0 | 0.00001 | 74 | 0 | 0 | 0 | 50 |
| 357 | 49878 | 1 | 0 | 0.00001 | 74 | 0 | 0 | 0 | 50 |
| 358 | 49878 | 1 | 0 | 0.00001 | 74 | 0 | 0 | 0 | 50 |
| 359 | 49878 | 1 | 0 | 0.00001 | 74 | 0 | 0 | 0 | 50 |
| 360 | 49878 | 1 | 0 | 0.00001 | 74 | 0 | 0 | 0 | 50 |
| 361 | 49877 | 2 | 0 | 0.00002 | 74 | 0 | 0 | 0 | 50 |
| 362 | 48437 | 1436 | 6 | 0.014515 | 73 | 1 | 0 | 0.006757 | 47.43 |
| 363 | 49878 | 1 | 0 | 0.00001 | 74 | 0 | 0 | 0 | 50 |
| 364 | 49876 | 3 | 0 | 0.00003 | 74 | 0 | 0 | 0 | 49.99 |
| 365 | 49877 | 2 | 0 | 0.00002 | 74 | 0 | 0 | 0 | 50 |
| 366 | 49878 | 1 | 0 | 0.00001 | 74 | 0 | 0 | 0 | 50 |
| 367 | 49867 | 6 | 0 | 0.00006 | 74 | 0 | 0 | 0 | 199.96 |
| 368 | 49878 | 1 | 0 | 0.00001 | 74 | 0 | 0 | 0 | 50 |
| 369 | 49875 | 4 | 0 | 0.00004 | 74 | 0 | 0 | 0 | 49.99 |
| 370 | 49878 | 1 | 0 | 0.00001 | 74 | 0 | 0 | 0 | 50 |
| 371 | 49878 | 1 | 0 | 0.00001 | 74 | 0 | 0 | 0 | 50 |
| 372 | 49878 | 1 | 0 | 0.00001 | 74 | 0 | 0 | 0 | 50 |
| 373 | 49877 | 2 | 0 | 0.00002 | 74 | 0 | 0 | 0 | 50 |
| 374 | 29726 | 17495 | 2658 | 0.228663 | 51 | 22 | 1 | 0.162162 | 18.26 |
| 375 | 49878 | 1 | 0 | 0.00001 | 74 | 0 | 0 | 0 | 50 |
| 376 | 49878 | 1 | 0 | 0.00001 | 74 | 0 | 0 | 0 | 50 |
| 377 | 49877 | 2 | 0 | 0.00002 | 74 | 0 | 0 | 0 | 50 |
| 378 | 49878 | 1 | 0 | 0.00001 | 74 | 0 | 0 | 0 | 50 |
| 379 | 49878 | 1 | 0 | 0.00001 | 74 | 0 | 0 | 0 | 50 |
| 380 | 49873 | 6 | 0 | 0.00006 | 74 | 0 | 0 | 0 | 49.99 |
| 381 | 49878 | 1 | 0 | 0.00001 | 74 | 0 | 0 | 0 | 50 |
| 382 | 49876 | 3 | 0 | 0.00003 | 74 | 0 | 0 | 0 | 30 |
| 383 | 49877 | 2 | 0 | 0.00002 | 74 | 0 | 0 | 0 | 30 |
| 384 | 49698 | 2 | 0 | 0.00002 | 74 | 0 | 0 | 0 | 30 |
| 385 | 49698 | 175 | 1 | 0.001774 | 74 | 0 | 0 | 0 | 29.81 |
| 386 | 49698 | 1 | 0 | 0.00001 | 74 | 0 | 0 | 0 | 30 |
| 387 | 49698 | 1 | 0 | 0.00001 | 74 | 0 | 0 | 0 | 30 |
| 388 | 49698 | 1 | 0 | 0.00001 | 74 | 0 | 0 | 0 | 30 |
| 389 | 49878 | 1 | 0 | 0.00001 | 74 | 0 | 0 | 0 | 30 |
| 390 | 49878 | 1 | 0 | 0.00001 | 74 | 0 | 0 | 0 | 30 |
| 391 | 49876 | 3 | 0 | 0.00003 | 74 | 0 | 0 | 0 | 30 |
| 392 | 49877 | 2 | 0 | 0.00002 | 74 | 0 | 0 | 0 | 30 |
| 393 | 49878 | 1 | 0 | 0.00001 | 74 | 0 | 0 | 0 | 30 |
| 394 | 49832 | 46 | 1 | 0.000481 | 74 | 0 | 0 | 0 | 29.95 |
| 395 | 49877 | 2 | 0 | 0.00002 | 74 | 0 | 0 | 0 | 30 |
| 396 | 0 | 1 | 49876 | 0.99999 | 0 | 0 | 74 | 1 | 30 |
| 397 | 49877 | 2 | 0 | 0.00002 | 74 | 0 | 0 | 0 | 30 |
| 398 | 49877 | 2 | 0 | 0.00002 | 74 | 0 | 0 | 0 | 30 |
| 399 | 49878 | 1 | 0 | 0.00001 | 74 | 0 | 0 | 0 | 30 |
| 400 | 49830 | 4 | 0 | 0.00004 | 74 | 0 | 0 | 0 | 30 |
| 401 | 49871 | 8 | 0 | 0.00008 | 74 | 0 | 0 | 0 | 29.99 |
| 402 | 49877 | 2 | 0 | 0.00002 | 74 | 0 | 0 | 0 | 30 |
| 403 | 49876 | 3 | 0 | 0.00003 | 74 | 0 | 0 | 0 | 30 |
| 404 | 49874 | 2 | 0 | 0.00002 | 74 | 0 | 0 | 0 | 30 |
| 405 | 49878 | 1 | 0 | 0.00001 | 74 | 0 | 0 | 0 | 30 |
| 406 | 49878 | 1 | 0 | 0.00001 | 74 | 0 | 0 | 0 | 30 |
| 407 | 49876 | 3 | 0 | 0.00003 | 74 | 0 | 0 | 0 | 30 |
| 408 | 49875 | 4 | 0 | 0.00004 | 74 | 0 | 0 | 0 | 30 |
| 409 | 24968 | 20457 | 4454 | 0.294362 | 41 | 28 | 5 | 0.256757 | 2.52 |
| 410 | 49878 | 1 | 0 | 0.00001 | 74 | 0 | 0 | 0 | 50 |
| 411 | 49875 | 4 | 0 | 0.00004 | 74 | 0 | 0 | 0 | 49.99 |
| 412 | 49874 | 5 | 0 | 0.00005 | 74 | 0 | 0 | 0 | 49.99 |
| 413 | 49877 | 2 | 0 | 0.00002 | 74 | 0 | 0 | 0 | 50 |
| 414 | 49868 | 1 | 0 | 0.00001 | 74 | 0 | 0 | 0 | 50 |
| 415 | 49877 | 2 | 0 | 0.00002 | 74 | 0 | 0 | 0 | 50 |
| 416 | 49877 | 2 | 0 | 0.00002 | 74 | 0 | 0 | 0 | 50 |
| 417 | 49877 | 2 | 0 | 0.00002 | 74 | 0 | 0 | 0 | 50 |
| 418 | 49870 | 9 | 0 | 0.00009 | 74 | 0 | 0 | 0 | 49.98 |
| 419 | 49878 | 1 | 0 | 0.00001 | 74 | 0 | 0 | 0 | 50 |
| 420 | 49878 | 1 | 0 | 0.00001 | 74 | 0 | 0 | 0 | 50 |
| 421 | 49878 | 1 | 0 | 0.00001 | 74 | 0 | 0 | 0 | 50 |
| 422 | 49877 | 1 | 0 | 0.00001 | 74 | 0 | 0 | 0 | 199.99 |
| 423 | 49877 | 1 | 0 | 0.00001 | 74 | 0 | 0 | 0 | 50 |
| 424 | 49877 | 1 | 0 | 0.00001 | 74 | 0 | 0 | 0 | 50 |
| 425 | 49875 | 3 | 0 | 0.00003 | 74 | 0 | 0 | 0 | 49.99 |
| 426 | 49877 | 1 | 0 | 0.00001 | 74 | 0 | 0 | 0 | 50 |
| 427 | 49873 | 5 | 0 | 0.00005 | 74 | 0 | 0 | 0 | 49.99 |
| 428 | 49877 | 1 | 0 | 0.00001 | 74 | 0 | 0 | 0 | 50 |
| 429 | 49876 | 2 | 0 | 0.00002 | 74 | 0 | 0 | 0 | 50 |
| 430 | 49878 | 1 | 0 | 0.00001 | 74 | 0 | 0 | 0 | 50 |
| 431 | 49878 | 1 | 0 | 0.00001 | 74 | 0 | 0 | 0 | 30 |
| 432 | 49878 | 1 | 0 | 0.00001 | 74 | 0 | 0 | 0 | 30 |
| 433 | 49878 | 1 | 0 | 0.00001 | 74 | 0 | 0 | 0 | 30 |
| 434 | 49878 | 1 | 0 | 0.00001 | 74 | 0 | 0 | 0 | 30 |
| 435 | 49870 | 9 | 0 | 0.00009 | 74 | 0 | 0 | 0 | 29.99 |
| 436 | 49807 | 71 | 1 | 0.000732 | 74 | 0 | 0 | 0 | 29.92 |
| 437 | 49873 | 5 | 0 | 0.00005 | 74 | 0 | 0 | 0 | 29.99 |
| 438 | 49857 | 21 | 0 | 0.000211 | 74 | 0 | 0 | 0 | 29.98 |
| 439 | 49774 | 98 | 2 | 0.001023 | 74 | 0 | 0 | 0 | 29.89 |
| 440 | 0 | 1 | 49872 | 0.99999 | 0 | 0 | 74 | 1 | 30 |
| 441 | 49872 | 1 | 0 | 0.00001 | 74 | 0 | 0 | 0 | 30 |
| 442 | 47405 | 2419 | 42 | 0.025097 | 71 | 3 | 0 | 0.02027 | 27.36 |
| 443 | 49381 | 36 | 0 | 0.000364 | 74 | 0 | 0 | 0 | 29.96 |
| 444 | 49834 | 1 | 0 | 0.00001 | 74 | 0 | 0 | 0 | 30 |
| 445 | 49780 | 68 | 1 | 0.000702 | 73 | 1 | 0 | 0.006757 | 29.92 |
| 446 | 49871 | 1 | 0 | 0.00001 | 74 | 0 | 0 | 0 | 30 |
| 447 | 49875 | 2 | 0 | 0.00002 | 74 | 0 | 0 | 0 | 30 |
| 448 | 49873 | 5 | 0 | 0.00005 | 74 | 0 | 0 | 0 | 29.99 |
| 449 | 49877 | 2 | 0 | 0.00002 | 74 | 0 | 0 | 0 | 30 |
| 450 | 49877 | 2 | 0 | 0.00002 | 74 | 0 | 0 | 0 | 30 |
| 451 | 49878 | 1 | 0 | 0.00001 | 74 | 0 | 0 | 0 | 50 |
| 452 | 49878 | 1 | 0 | 0.00001 | 74 | 0 | 0 | 0 | 50 |
| 453 | 49878 | 1 | 0 | 0.00001 | 74 | 0 | 0 | 0 | 50 |
| 454 | 49877 | 2 | 0 | 0.00002 | 74 | 0 | 0 | 0 | 50 |
| 455 | 49874 | 5 | 0 | 0.00005 | 74 | 0 | 0 | 0 | 49.99 |
| 456 | 49877 | 1 | 0 | 0.00001 | 74 | 0 | 0 | 0 | 50 |
| 457 | 49875 | 3 | 0 | 0.00003 | 74 | 0 | 0 | 0 | 49.99 |
| 458 | 49876 | 1 | 0 | 0.00001 | 74 | 0 | 0 | 0 | 100 |
| 459 | 49878 | 1 | 0 | 0.00001 | 74 | 0 | 0 | 0 | 50 |
| 460 | 49845 | 17 | 0 | 0.00017 | 74 | 0 | 0 | 0 | 299.82 |
| 461 | 49878 | 1 | 0 | 0.00001 | 74 | 0 | 0 | 0 | 50 |
| 462 | 49878 | 1 | 0 | 0.00001 | 74 | 0 | 0 | 0 | 50 |
| 463 | 49878 | 1 | 0 | 0.00001 | 74 | 0 | 0 | 0 | 50 |
| 464 | 49878 | 1 | 0 | 0.00001 | 74 | 0 | 0 | 0 | 50 |
| 465 | 49878 | 1 | 0 | 0.00001 | 74 | 0 | 0 | 0 | 50 |
| 466 | 49878 | 1 | 0 | 0.00001 | 74 | 0 | 0 | 0 | 30 |
| 467 | 49857 | 15 | 0 | 0.00015 | 74 | 0 | 0 | 0 | 29.98 |
| 468 | 48631 | 1213 | 9 | 0.012346 | 73 | 1 | 0 | 0.006757 | 28.68 |
| 469 | 49733 | 1 | 0 | 0.00001 | 74 | 0 | 0 | 0 | 30 |
| 470 | 49733 | 1 | 0 | 0.00001 | 74 | 0 | 0 | 0 | 30 |
| 471 | 49834 | 1 | 0 | 0.00001 | 74 | 0 | 0 | 0 | 30 |
| 472 | 49848 | 1 | 0 | 0.00001 | 74 | 0 | 0 | 0 | 30 |
| 473 | 49856 | 6 | 0 | 0.00006 | 74 | 0 | 0 | 0 | 29.99 |
| 474 | 49850 | 17 | 0 | 0.00017 | 74 | 0 | 0 | 0 | 29.98 |
| 475 | 49878 | 1 | 0 | 0.00001 | 74 | 0 | 0 | 0 | 30 |
| 476 | 49878 | 1 | 0 | 0.00001 | 74 | 0 | 0 | 0 | 30 |
| 477 | 49636 | 243 | 0 | 0.002436 | 74 | 0 | 0 | 0 | 29.74 |
| 478 | 49878 | 1 | 0 | 0.00001 | 74 | 0 | 0 | 0 | 30 |
| 479 | 49878 | 1 | 0 | 0.00001 | 74 | 0 | 0 | 0 | 30 |
| 480 | 49878 | 1 | 0 | 0.00001 | 74 | 0 | 0 | 0 | 30 |
| 481 | 49864 | 15 | 0 | 0.00015 | 74 | 0 | 0 | 0 | 29.98 |
| 482 | 49875 | 4 | 0 | 0.00004 | 74 | 0 | 0 | 0 | 30 |
| 483 | 49876 | 3 | 0 | 0.00003 | 74 | 0 | 0 | 0 | 30 |
| 484 | 49878 | 1 | 0 | 0.00001 | 74 | 0 | 0 | 0 | 30 |
| 485 | 49867 | 12 | 0 | 0.00012 | 74 | 0 | 0 | 0 | 29.99 |
| 486 | 49877 | 1 | 0 | 0.00001 | 74 | 0 | 0 | 0 | 30 |
| 487 | 49877 | 1 | 0 | 0.00001 | 74 | 0 | 0 | 0 | 30 |
| 488 | 49878 | 1 | 0 | 0.00001 | 74 | 0 | 0 | 0 | 30 |
| 489 | 49878 | 1 | 0 | 0.00001 | 74 | 0 | 0 | 0 | 50 |
| 490 | 49878 | 1 | 0 | 0.00001 | 74 | 0 | 0 | 0 | 50 |
| 491 | 49878 | 1 | 0 | 0.00001 | 74 | 0 | 0 | 0 | 50 |
| 492 | 42095 | 7392 | 392 | 0.081958 | 61 | 12 | 1 | 0.094595 | 36.45 |
| 493 | 48949 | 925 | 5 | 0.009373 | 73 | 1 | 0 | 0.006757 | 48.33 |
| 494 | 49857 | 5 | 0 | 0.00005 | 74 | 0 | 0 | 0 | 99.98 |
| 495 | 49878 | 1 | 0 | 0.00001 | 74 | 0 | 0 | 0 | 50 |
| 496 | 49875 | 4 | 0 | 0.00004 | 74 | 0 | 0 | 0 | 49.99 |
| 497 | 49877 | 2 | 0 | 0.00002 | 74 | 0 | 0 | 0 | 50 |
| 498 | 49877 | 2 | 0 | 0.00002 | 74 | 0 | 0 | 0 | 50 |
| 499 | 49862 | 17 | 0 | 0.00017 | 74 | 0 | 0 | 0 | 49.97 |
| 500 | 49873 | 6 | 0 | 0.00006 | 74 | 0 | 0 | 0 | 49.99 |
| 501 | 48487 | 1377 | 15 | 0.014104 | 72 | 2 | 0 | 0.013514 | 47.5 |
| 502 | 49878 | 1 | 0 | 0.00001 | 74 | 0 | 0 | 0 | 199.99 |
| 503 | 49878 | 1 | 0 | 0.00001 | 74 | 0 | 0 | 0 | 50 |
| 504 | 49878 | 1 | 0 | 0.00001 | 74 | 0 | 0 | 0 | 50 |
| 505 | 49877 | 2 | 0 | 0.00002 | 74 | 0 | 0 | 0 | 50 |
| 506 | 49878 | 1 | 0 | 0.00001 | 74 | 0 | 0 | 0 | 50 |
| 507 | 49870 | 9 | 0 | 0.00009 | 74 | 0 | 0 | 0 | 49.98 |
| 508 | 49870 | 3 | 0 | 0.00003 | 74 | 0 | 0 | 0 | 199.98 |
| 509 | 49870 | 2 | 0 | 0.00002 | 74 | 0 | 0 | 0 | 299.98 |
| 510 | 49872 | 1 | 0 | 0.00001 | 74 | 0 | 0 | 0 | 50 |
| 511 | 49868 | 11 | 0 | 0.00011 | 74 | 0 | 0 | 0 | 49.98 |
| 512 | 49878 | 1 | 0 | 0.00001 | 74 | 0 | 0 | 0 | 50 |
| 513 | 49876 | 3 | 0 | 0.00003 | 74 | 0 | 0 | 0 | 49.99 |
| 514 | 49878 | 1 | 0 | 0.00001 | 74 | 0 | 0 | 0 | 50 |
| 515 | 49878 | 1 | 0 | 0.00001 | 74 | 0 | 0 | 0 | 50 |
| 516 | 49878 | 1 | 0 | 0.00001 | 74 | 0 | 0 | 0 | 50 |
| 517 | 49833 | 46 | 0 | 0.000461 | 74 | 0 | 0 | 0 | 49.92 |
| 518 | 49878 | 1 | 0 | 0.00001 | 74 | 0 | 0 | 0 | 50 |
| 519 | 49875 | 4 | 0 | 0.00004 | 74 | 0 | 0 | 0 | 49.99 |
| 520 | 49877 | 2 | 0 | 0.00002 | 74 | 0 | 0 | 0 | 50 |
| 521 | 49877 | 2 | 0 | 0.00002 | 74 | 0 | 0 | 0 | 50 |
| 522 | 49878 | 1 | 0 | 0.00001 | 74 | 0 | 0 | 0 | 50 |
| 523 | 49877 | 2 | 0 | 0.00002 | 74 | 0 | 0 | 0 | 50 |
| 524 | 49846 | 33 | 0 | 0.000331 | 74 | 0 | 0 | 0 | 49.94 |
| 525 | 49876 | 3 | 0 | 0.00003 | 74 | 0 | 0 | 0 | 49.99 |
| 526 | 49875 | 4 | 0 | 0.00004 | 74 | 0 | 0 | 0 | 49.99 |
| 527 | 49868 | 11 | 0 | 0.00011 | 74 | 0 | 0 | 0 | 199.92 |
| 528 | 49611 | 267 | 1 | 0.002697 | 74 | 0 | 0 | 0 | 49.52 |
| 529 | 49872 | 1 | 0 | 0.00001 | 74 | 0 | 0 | 0 | 50 |
| 530 | 49876 | 3 | 0 | 0.00003 | 74 | 0 | 0 | 0 | 49.99 |
| 531 | 49878 | 1 | 0 | 0.00001 | 74 | 0 | 0 | 0 | 50 |
| 532 | 49876 | 3 | 0 | 0.00003 | 74 | 0 | 0 | 0 | 49.99 |
| 533 | 49877 | 2 | 0 | 0.00002 | 74 | 0 | 0 | 0 | 50 |
| 534 | 49860 | 19 | 0 | 0.00019 | 74 | 0 | 0 | 0 | 49.97 |
| 535 | 49878 | 1 | 0 | 0.00001 | 74 | 0 | 0 | 0 | 50 |
| 536 | 49878 | 1 | 0 | 0.00001 | 74 | 0 | 0 | 0 | 50 |
| 537 | 49877 | 2 | 0 | 0.00002 | 74 | 0 | 0 | 0 | 50 |
| 538 | 49878 | 1 | 0 | 0.00001 | 74 | 0 | 0 | 0 | 50 |
| 539 | 49875 | 4 | 0 | 0.00004 | 74 | 0 | 0 | 0 | 49.99 |
| 540 | 49878 | 1 | 0 | 0.00001 | 74 | 0 | 0 | 0 | 50 |
| 541 | 49877 | 2 | 0 | 0.00002 | 74 | 0 | 0 | 0 | 50 |
| 542 | 49877 | 2 | 0 | 0.00002 | 74 | 0 | 0 | 0 | 50 |
| 543 | 49869 | 10 | 0 | 0.0001 | 74 | 0 | 0 | 0 | 29.99 |
| 544 | 49875 | 1 | 0 | 0.00001 | 74 | 0 | 0 | 0 | 30 |
| 545 | 49868 | 11 | 0 | 0.00011 | 74 | 0 | 0 | 0 | 29.99 |
| 546 | 49875 | 2 | 0 | 0.00002 | 74 | 0 | 0 | 0 | 30 |
| 547 | 49875 | 1 | 0 | 0.00001 | 74 | 0 | 0 | 0 | 30 |
| 548 | 49870 | 1 | 0 | 0.00001 | 74 | 0 | 0 | 0 | 30 |
| 549 | 49870 | 1 | 0 | 0.00001 | 74 | 0 | 0 | 0 | 30 |
| 550 | 49867 | 4 | 0 | 0.00004 | 74 | 0 | 0 | 0 | 30 |
| 551 | 49865 | 1 | 0 | 0.00001 | 74 | 0 | 0 | 0 | 30 |
| 552 | 49772 | 91 | 0 | 0.000913 | 74 | 0 | 0 | 0 | 29.9 |
| 553 | 49804 | 1 | 0 | 0.00001 | 73 | 0 | 0 | 0 | 30 |
| 554 | 49759 | 1 | 0 | 0.00001 | 73 | 0 | 0 | 0 | 30 |
| 555 | 49625 | 2 | 0 | 0.00002 | 73 | 0 | 0 | 0 | 30 |
| 556 | 49620 | 1 | 0 | 0.00001 | 73 | 0 | 0 | 0 | 30 |
| 557 | 49800 | 1 | 0 | 0.00001 | 74 | 0 | 0 | 0 | 30 |
| 558 | 49850 | 1 | 0 | 0.00001 | 74 | 0 | 0 | 0 | 30 |
| 559 | 49847 | 5 | 0 | 0.00005 | 74 | 0 | 0 | 0 | 29.99 |
| 560 | 49866 | 3 | 0 | 0.00003 | 74 | 0 | 0 | 0 | 30 |
| 561 | 49877 | 2 | 0 | 0.00002 | 74 | 0 | 0 | 0 | 30 |
| 562 | 49873 | 6 | 0 | 0.00006 | 74 | 0 | 0 | 0 | 49.99 |
| 563 | 49876 | 3 | 0 | 0.00003 | 74 | 0 | 0 | 0 | 49.99 |
| 564 | 49878 | 1 | 0 | 0.00001 | 74 | 0 | 0 | 0 | 50 |
| 565 | 49877 | 2 | 0 | 0.00002 | 74 | 0 | 0 | 0 | 50 |
| 566 | 49878 | 1 | 0 | 0.00001 | 74 | 0 | 0 | 0 | 50 |
| 567 | 49872 | 3 | 0 | 0.00003 | 74 | 0 | 0 | 0 | 30 |
| 568 | 49842 | 1 | 0 | 0.00001 | 74 | 0 | 0 | 0 | 30 |
| 569 | 49860 | 1 | 0 | 0.00001 | 74 | 0 | 0 | 0 | 30 |
| 570 | 49778 | 3 | 0 | 0.00003 | 74 | 0 | 0 | 0 | 30 |
| 571 | 49347 | 1 | 0 | 0.00001 | 74 | 0 | 0 | 0 | 30 |
| 572 | 49177 | 13 | 0 | 0.000132 | 74 | 0 | 0 | 0 | 29.99 |
| 573 | 49167 | 1 | 0 | 0.00001 | 74 | 0 | 0 | 0 | 30 |
| 574 | 49878 | 1 | 0 | 0.00001 | 74 | 0 | 0 | 0 | 30 |
| 575 | 49878 | 1 | 0 | 0.00001 | 74 | 0 | 0 | 0 | 30 |
| 576 | 49874 | 1 | 0 | 0.00001 | 74 | 0 | 0 | 0 | 30 |
| 577 | 49853 | 26 | 0 | 0.000261 | 74 | 0 | 0 | 0 | 29.97 |
| 578 | 49875 | 4 | 0 | 0.00004 | 74 | 0 | 0 | 0 | 30 |
| 579 | 49878 | 1 | 0 | 0.00001 | 74 | 0 | 0 | 0 | 30 |
| 580 | 49878 | 1 | 0 | 0.00001 | 74 | 0 | 0 | 0 | 30 |
| 581 | 49878 | 1 | 0 | 0.00001 | 74 | 0 | 0 | 0 | 30 |
| 582 | 49878 | 1 | 0 | 0.00001 | 74 | 0 | 0 | 0 | 30 |
| 583 | 49847 | 32 | 0 | 0.000321 | 74 | 0 | 0 | 0 | 29.97 |
| 584 | 49864 | 15 | 0 | 0.00015 | 74 | 0 | 0 | 0 | 29.98 |
| 585 | 49878 | 1 | 0 | 0.00001 | 74 | 0 | 0 | 0 | 30 |
| 586 | 49878 | 1 | 0 | 0.00001 | 74 | 0 | 0 | 0 | 30 |
| 587 | 49878 | 1 | 0 | 0.00001 | 74 | 0 | 0 | 0 | 30 |
| 588 | 49878 | 1 | 0 | 0.00001 | 74 | 0 | 0 | 0 | 30 |
| 589 | 49835 | 44 | 0 | 0.000441 | 74 | 0 | 0 | 0 | 29.95 |
| 590 | 49878 | 1 | 0 | 0.00001 | 74 | 0 | 0 | 0 | 30 |
| 591 | 49877 | 2 | 0 | 0.00002 | 74 | 0 | 0 | 0 | 30 |
| 592 | 49875 | 4 | 0 | 0.00004 | 74 | 0 | 0 | 0 | 30 |
| 593 | 49877 | 2 | 0 | 0.00002 | 74 | 0 | 0 | 0 | 30 |
| 594 | 49878 | 1 | 0 | 0.00001 | 74 | 0 | 0 | 0 | 30 |
| 595 | 49878 | 1 | 0 | 0.00001 | 74 | 0 | 0 | 0 | 30 |
| 596 | 49878 | 1 | 0 | 0.00001 | 74 | 0 | 0 | 0 | 30 |
| 597 | 49876 | 3 | 0 | 0.00003 | 74 | 0 | 0 | 0 | 30 |
| 598 | 49878 | 1 | 0 | 0.00001 | 74 | 0 | 0 | 0 | 30 |
| 599 | 49878 | 1 | 0 | 0.00001 | 74 | 0 | 0 | 0 | 30 |
| 600 | 49877 | 2 | 0 | 0.00002 | 74 | 0 | 0 | 0 | 30 |
| 601 | 49878 | 1 | 0 | 0.00001 | 74 | 0 | 0 | 0 | 30 |
| 602 | 49878 | 1 | 0 | 0.00001 | 74 | 0 | 0 | 0 | 30 |
| 603 | 49876 | 3 | 0 | 0.00003 | 74 | 0 | 0 | 0 | 30 |
| 604 | 49878 | 1 | 0 | 0.00001 | 74 | 0 | 0 | 0 | 30 |
| 605 | 49876 | 3 | 0 | 0.00003 | 74 | 0 | 0 | 0 | 30 |
| 606 | 49878 | 1 | 0 | 0.00001 | 74 | 0 | 0 | 0 | 30 |
| 607 | 49868 | 11 | 0 | 0.00011 | 74 | 0 | 0 | 0 | 29.99 |
| 608 | 49878 | 1 | 0 | 0.00001 | 74 | 0 | 0 | 0 | 30 |
| 609 | 49878 | 1 | 0 | 0.00001 | 74 | 0 | 0 | 0 | 30 |
| 610 | 49876 | 3 | 0 | 0.00003 | 74 | 0 | 0 | 0 | 30 |
| 611 | 49862 | 17 | 0 | 0.00017 | 74 | 0 | 0 | 0 | 29.98 |
| 612 | 49878 | 1 | 0 | 0.00001 | 74 | 0 | 0 | 0 | 30 |
| 613 | 49878 | 1 | 0 | 0.00001 | 74 | 0 | 0 | 0 | 30 |
| 614 | 49878 | 1 | 0 | 0.00001 | 74 | 0 | 0 | 0 | 30 |
| 615 | 49850 | 29 | 0 | 0.000291 | 74 | 0 | 0 | 0 | 29.97 |
| 616 | 49873 | 5 | 0 | 0.00005 | 74 | 0 | 0 | 0 | 29.99 |
| 617 | 49875 | 1 | 0 | 0.00001 | 74 | 0 | 0 | 0 | 30 |
| 618 | 49878 | 1 | 0 | 0.00001 | 74 | 0 | 0 | 0 | 30 |
| 619 | 49878 | 1 | 0 | 0.00001 | 74 | 0 | 0 | 0 | 30 |
| 620 | 49876 | 3 | 0 | 0.00003 | 74 | 0 | 0 | 0 | 30 |
| 621 | 49877 | 2 | 0 | 0.00002 | 74 | 0 | 0 | 0 | 30 |
| 622 | 49876 | 3 | 0 | 0.00003 | 74 | 0 | 0 | 0 | 30 |
| 623 | 49704 | 175 | 0 | 0.001754 | 74 | 0 | 0 | 0 | 29.81 |
| 624 | 49878 | 1 | 0 | 0.00001 | 74 | 0 | 0 | 0 | 30 |
| 625 | 49878 | 1 | 0 | 0.00001 | 74 | 0 | 0 | 0 | 30 |
| 626 | 49878 | 1 | 0 | 0.00001 | 74 | 0 | 0 | 0 | 30 |
| 627 | 49878 | 1 | 0 | 0.00001 | 74 | 0 | 0 | 0 | 30 |
| 628 | 49878 | 1 | 0 | 0.00001 | 74 | 0 | 0 | 0 | 30 |
| 629 | 49874 | 5 | 0 | 0.00005 | 74 | 0 | 0 | 0 | 29.99 |
| 630 | 49878 | 1 | 0 | 0.00001 | 74 | 0 | 0 | 0 | 30 |
| 631 | 49875 | 4 | 0 | 0.00004 | 74 | 0 | 0 | 0 | 30 |
| 632 | 49878 | 1 | 0 | 0.00001 | 74 | 0 | 0 | 0 | 30 |
| 633 | 49872 | 7 | 0 | 0.00007 | 74 | 0 | 0 | 0 | 49.99 |
| 634 | 49877 | 1 | 0 | 0.00001 | 74 | 0 | 0 | 0 | 199.99 |
| 635 | 49877 | 1 | 0 | 0.00001 | 74 | 0 | 0 | 0 | 199.99 |
| 636 | 49873 | 6 | 0 | 0.00006 | 74 | 0 | 0 | 0 | 49.99 |
| 637 | 49869 | 10 | 0 | 0.0001 | 74 | 0 | 0 | 0 | 49.98 |
| 638 | 49877 | 2 | 0 | 0.00002 | 74 | 0 | 0 | 0 | 50 |
| 639 | 18275 | 23765 | 7839 | 0.395387 | 24 | 35 | 15 | 0.439189 | 6.97 |
| 640 | 48676 | 1194 | 9 | 0.012149 | 74 | 0 | 0 | 0 | 47.84 |
| 641 | 49878 | 1 | 0 | 0.00001 | 74 | 0 | 0 | 0 | 199.99 |
| 642 | 49878 | 1 | 0 | 0.00001 | 74 | 0 | 0 | 0 | 50 |
| 643 | 49877 | 2 | 0 | 0.00002 | 74 | 0 | 0 | 0 | 50 |
| 644 | 49877 | 2 | 0 | 0.00002 | 74 | 0 | 0 | 0 | 50 |
| 645 | 49877 | 2 | 0 | 0.00002 | 74 | 0 | 0 | 0 | 50 |
| 646 | 49872 | 1 | 0 | 0.00001 | 74 | 0 | 0 | 0 | 50 |
| 647 | 49878 | 1 | 0 | 0.00001 | 74 | 0 | 0 | 0 | 50 |
| 648 | 49878 | 1 | 0 | 0.00001 | 74 | 0 | 0 | 0 | 50 |
| 649 | 49876 | 3 | 0 | 0.00003 | 74 | 0 | 0 | 0 | 49.99 |
| 650 | 49878 | 1 | 0 | 0.00001 | 74 | 0 | 0 | 0 | 50 |
| 651 | 49878 | 1 | 0 | 0.00001 | 74 | 0 | 0 | 0 | 50 |
| 652 | 49878 | 1 | 0 | 0.00001 | 74 | 0 | 0 | 0 | 50 |
| 653 | 49877 | 1 | 0 | 0.00001 | 74 | 0 | 0 | 0 | 50 |
| 654 | 49878 | 1 | 0 | 0.00001 | 74 | 0 | 0 | 0 | 50 |
| 655 | 49874 | 2 | 0 | 0.00002 | 74 | 0 | 0 | 0 | 50 |
| 656 | 49878 | 1 | 0 | 0.00001 | 74 | 0 | 0 | 0 | 50 |
| 657 | 49878 | 1 | 0 | 0.00001 | 74 | 0 | 0 | 0 | 50 |
| 658 | 49878 | 1 | 0 | 0.00001 | 74 | 0 | 0 | 0 | 50 |

